# Association of Sedentary and Physical Activity Behaviors with Body Composition: a Genome-Wide Association and Mendelian Randomization Study

**DOI:** 10.1101/2022.04.20.22274068

**Authors:** Ferris Ramadan, Jennifer W. Bea, David O. Garcia, Katherine Ellingson, Robert A. Canales, David A. Raichlen, Yann C. Klimentidis

## Abstract

**Objectives:** Studies suggest that body composition can be independently improved through physical activity (PA). We performed a Mendelian randomization (MR) study to test the incremental benefits of sedentary behavior and various physical activity (PA) exposures on body composition outcomes as assessed by anthropometric indices, lean body mass (LBM) (*kg*), body fat (*%*), and visceral adipose tissue (VAT) (*kg*).

**Methods:** Genetic instruments were identified for both self-reported and accelerometer-measured sedentary behavior and PA. Outcomes included anthropometric and dual-energy X-ray absorptiometry measures of adiposity, extracted from the UK Biobank and the largest available consortia. Multivariable MR (MVMR) included educational attainment as a covariate to address potential confounding. Sensitivity analyses were evaluated for weak instrument bias and pleiotropic effects.

**Results:** We did not identify consistent associations between genetically-predicted self-reported and accelerometer-measured sedentary behavior and body composition outcomes. All analyses for self-reported moderate PA were null for body composition outcomes. Genetically-predicted PA at higher intensities was protective against VAT in MR and MVMR analyses of both accelerometer-measured vigorous PA (MVMR *β* = -0.15, 95% Confidence Interval (CI): -0.24, -0.07, p<0.001) and self-reported participation in strenuous sports or other exercises (MVMR *β* = -0.27, 95%CI: -0.52, -0.01, p=0.034) was robust across several sensitivity analyses.

**Conclusions:** We did not identify evidence of a causal relationship between genetically-predicted PA and body composition, with the exception of a putatively protective effect of higher-intensity PA on VAT. Protective effects of PA against VAT may support prior evidence of biological pathways through which PA decreases risk of downstream cardiometabolic diseases.

## INTRODUCTION

Obesity has reached critical levels, and continues to exacerbate pervasive public health concerns, including cardiovascular diseases, cancers, type-2 diabetes, mental health and discrimination.(1) This high prevalence has been ascribed to an increasingly obesogenic environment, characterized by reduced physical activity (PA), increased occupational and recreational sedentary activities, and dietary options inundated by highly palatable energy-dense foods.(2) In an effort to reduce body fat, weight management programs have principally targeted both a reduction in energy intake and increased PA levels. Although PA offers substantial benefits to overall health,(3) these efforts have largely fallen short in attenuating obesity rates.(4–6)

Interventions have examined the putative effect of PA on various body composition measures, such as body mass index (BMI), waist circumference (WC), waist-to-hip ratio (WHR) and total body fat percent (%TBF). Combined diet and exercise programs are optimal for improving body composition versus diet-only programs;(7) however, PA alone inconsistently protects against clinically-significant weight gain.(8) Previous studies examining the effect of PA on lean body mass (LBM) – an aggregate body mass measurement excluding fat – have similarly yielded mixed outcomes.(9–11) Current evidence suggests that at least 150 minutes/week spent in moderate PA offers the greatest protection against weight gain.(12,13) Although there is limited evidence indicating a dose-response relationship between PA and weight gain, PA programs prescribing higher-intensity exercise have resulted in greater mass reduction, compared to lower-intensity programs.(14,15) Of particular importance for improving body composition is reducing visceral adipose tissue (VAT), which is a stronger predictor of mortality compared to non-specific anthropometric and subcutaneous fat measures.(16) Observational studies have supported significant benefits of PA on VAT reduction, independently of diet.(17–20) High-intensity interval training interventions have further demonstrated VAT reductions with similar results to mixed diet and PA programs.(21) Understanding the effect of PA on VAT thus holds particular value in attenuating the risk of associated downstream diseases.

Genome wide association studies (GWAS) have identified multiple genetic variants associated with PA propensity (22,23) and body composition measures.(24–27) Here, we use Mendelian randomization (MR) to evaluate the relationship of sedentary behavior and PA intensity with body composition. MR is a form of instrumental variable analysis that leverages genetic variants from GWAS to infer causality between an exposure and a given outcome.(28) The assignment of genetic variants during gamete formation confers a lifelong exposure for individuals randomized to a given allelic variant. Unlike exposures measured at the phenotype level, genetic variants are not typically associated with a multitude of behavioral, social, or physiological factors that may confound an epidemiological association.(29) MR may thus avoid many of the pitfalls seen in previous observational studies, including confounding (30) and reverse causation.(31) In this study, we use MR to consider sedentary behavior, multiple PA exposures and multiple body composition outcomes which may provide a more granular understanding of the benefit, if any, of activity levels on body composition.

## METHODS

### Study design

We performed MR and multivariable MR (MVMR) analyses to evaluate the dose-response relationship between sedentary behavior, PA and 1) whole LBM, 2) BMI, 3) WC, 4) WHR, 5) %TBF and 6) VAT (Figure 1). Exposure and outcome instruments were derived from non-overlapping samples to facilitate application of a two-sample MR study, which is less prone to weak instrument bias compared to one-sample MR. Both self-reported and accelerometer-measured exposures were used to evaluate PA associations with body composition measures. Self-reported and fraction acceleration thresholds were selected based on metabolic equivalent of tasks (METs), including sedentary behavior (≤1.5 METs), moderate PA (3.0–5.9 METs), moderate-to-vigorous PA (MVPA) (≥3.0 METs), and vigorous PA (≥6.0 METs). We did not consider the light PA phenotype due to the limited measurement sensitivity of these measurements. Self-reported strenuous sports or other exercises (SSOE) and overall acceleration measures were further included in analyses due to evidence suggesting strong genetic markers for these exposures.(22) MVMR analyses included educational attainment as a covariate, as described below. This study is reported according to the strengthening the reporting of observational studies in epidemiology using mendelian randomization (STROBE-MR) guidelines.(32)

**Figure 1.**
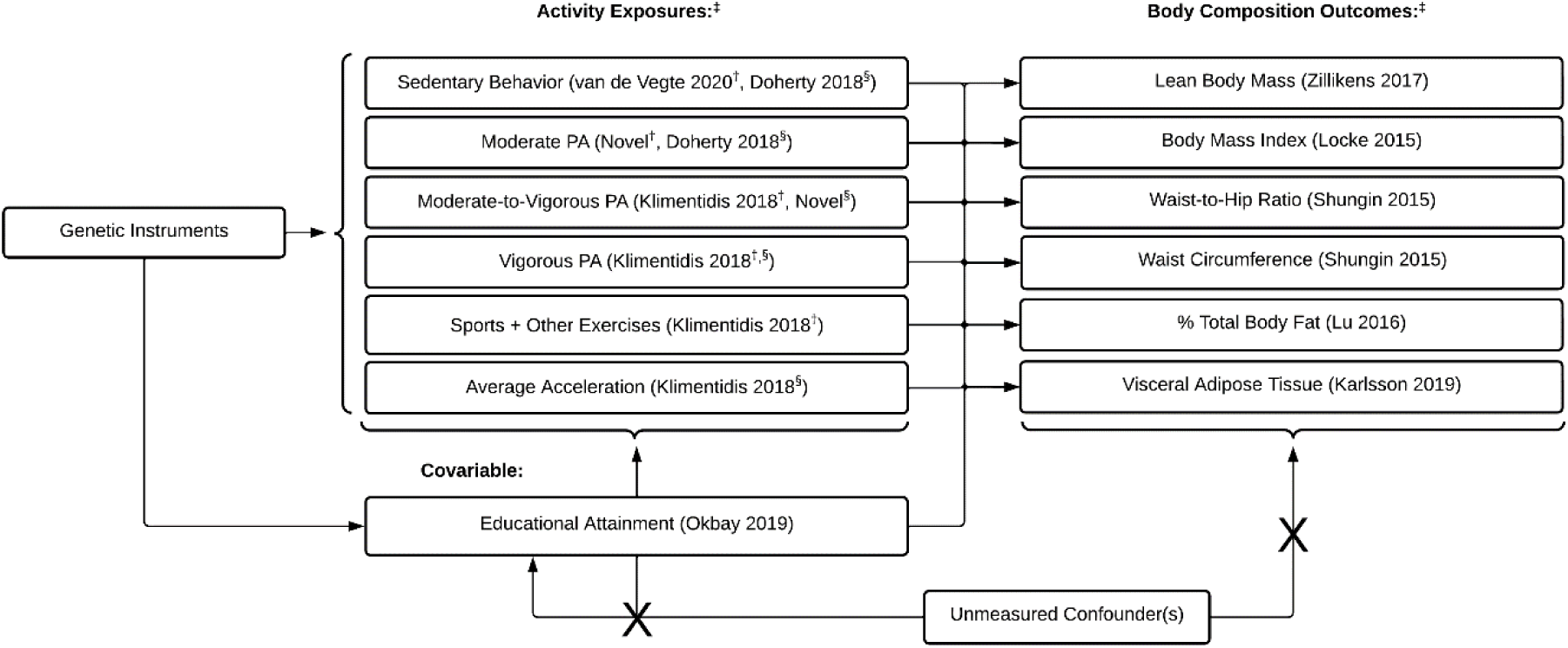
Sedentary Behavior and Physical Activity Exposures and Body Composition Outcomes Evaluated in Mendelian Randomization Analyses. Abbreviations: PA, Physical Activity. Note: ^‡^ GWAS author and publication year are reported adjacent to exposure and outcome phenotypes where applicable; ^†^ Self-reported exposure; ^§^ Accelerometer measured exposure

Genetic instruments were identified from GWAS from the UK Biobank (UKB), GEnetic Factors for OSteoporosis (GEFOS) consortium (33) and the Genetic Investigation of ANthropometric Traits (GIANT) consortium (25). The UKB is an ongoing prospective cohort study in the United Kingdom. Approximately a half-million adults, aged 37 – 73, have been enrolled and provided genotype and phenotype data to investigators. Neither patients nor the public were involved in the design, conduct, reporting, or dissemination of this research. Ethical approval of the UKB was provided by the Northwest Multicentre Research Ethics Committee, the National Information Governance Board for Health & Social Care, and the Community Health Index Advisory Group (IRAS project ID: 299116).

### Exposure Measures

Self-reported PA was previously captured for over 377 000 UKB participants via in-person questionnaires at the baseline exam (2006-2010). Self-reported total sedentary behavior was not measured directly in the UKB. Therefore, we used a combined proxy measure of sedentary behaviors, including average self-reported hours of leisure television watching, leisure computer use and driving time (n=422 218).(34) These behaviors show higher validity than total sedentary behavior and are easier to recall.(35)

Self-reported light PA was not included in analyses due to low specificity of available UKB phenotypes. Self-reported moderate and vigorous PA were captured directly in the UKB. MVPA was calculated using the International Physical Activity Questionnaire scoring protocol by taking the sum of total minutes/week of moderate PA multiplied by four and the total number of vigorous PA minutes/week multiplied by eight.(36,37) A summary of these phenotypes and quality control procedures have been previously described by Klimentidis et al.(22)

Acceleration data in the UKB were captured for over 90 000 participants over a one-week duration with the Axivity AX3 wrist-worn triaxial accelerometer.(23) Data were calibrated, resampled, and summarized using previously outlined procedures.(38) Accelerometer-derived PA can be converted to METs using fraction of acceleration time, measured in 5-second epochs of milligravities (*mg*). Sedentary and moderate PA METs were identified by Doherty et al. (23) on accelerometer data using predicted behavior states from a machine-learning model trained on free-living participants. We did not derive a light PA phenotype due to limited accelerometer sensitivity at lower-level accelerations, which may have introduced ambiguity in differentiating between activity states. MVPA was defined as the fraction of accelerations above 100*mg*.(39) Vigorous PA was defined as the fraction of accelerations above 425*mg*.(39) In summary, accelerometer-measured sedentary and moderate PA phenotypes were isolated using predicted behavior states, while MVPA and vigorous PA phenotypes were established from acceleration cut-points corresponding to METs.

Summary statistics were previously unavailable for self-reported moderate PA and accelerometer-measured PA. We conducted GWAS on 360 911 participants with self-reported moderate PA, and 97 737 participants with accelerometer-measured MVPA using the BOLT-LMM v2.3.6 software tool.(40) Briefly, participants were excluded based on >5% missing rates, unusually high heterozygosity, and a mismatch between self-reported and genetically inferred sex. To limit confounding from population-level stratification, only self-reported European participants were included in GWAS. SNPs with high missingness (>1.5%), genetic versus self-reported sex mismatches, low minor allele frequency (<0.1%), and low imputation quality were excluded from analyses.

In all models, we controlled for covariates including genotyping chip, center, age-squared, age, sex, age-sex interaction, and the first 10 principal components to correct for population stratification and genotyping array. Educational attainment was collected from multiple cohorts as a continuous measure of years of school competed (n=293 723).(41) We previously identified strong genetic correlations of education with PA.(22,42) MVMR analyses included educational attainment as a covariate to minimize potential confounding.

### Outcome Measures

Outcome measurements were extracted from non-overlapping studies. The LBM phenotype used in the GEFOS consortium was measured in kilograms (*kg)* using bioimpedance analysis (BIA) or dual energy X-ray absorptiometry (DEXA) (n=38 292).(24) Despite lower specificity from BIA measurements, BIA and DEXA show strong agreement and can be used interchangeably at the population level.(43) Summary statistics of GWAS (not including the UKB) for anthropometric outcomes including BMI (*kg/m*^2^) (n=249 796), WC adjusted for BMI (centimeters), and WHR adjusted for BMI (n=224 459) were obtained from the GIANT consortium.(25) We also used a GWAS meta-analysis that did not overlap with the UKB to extract summary statistics for %TBF (n=89 297).(26) The %TBF phenotype was measured with either BIA or DEXA. Large-scale GWAS of VAT has been limited, however, due to the need for advanced imaging phenotypes on large numbers of individuals to reveal significant loci. To reach a sample size that would yield greater genetic signal, we applied a recently developed non-linear VAT prediction model developed by Karlsson et al.(27) Briefly, the model was trained on 4 000 UKB participants with DEXA-measured VAT (*kg*). BIA measures on the other UKB participants were used as the primary predictive features within models. Despite a reduced sensitivity, the consistent relationship between BIA-derived measures and measured VAT suggests heightened model performance, and indicated strong predictive capacity (coefficient of determination (*r*^2^) = 0.76).(27) We performed GWAS for VAT on a subset of non-overlapping UKB participants including those without self-reported PA (n=56 908), and accelerometer measurements (n=323 769) (Table S1).

### Instrument Selection

We selected single nucleotide polymorphisms (SNPs) at p<5×10^−6^ for exposures. SNPs in linkage disequilibrium with the top SNP were removed to account for correlated genetic variants (*r*^2^<0.001). Proxies for genetic variants missing in outcome GWAS were identified using the 1 000 Genomes European sample data. Alleles were harmonized to ensure that the same allele was being referenced for exposure and outcome effect sizes. We attempted to infer the positive strand allele using allele frequencies for palindromic SNPs. We did not manually “prune” for genetic variants with suspected associations with outcome phenotypes as it risks the removal of only IVs with a directional effect.(44) We were unable to identify an adequate number of SNPs for accelerometer-measured moderate PA exposures, and for sedentary behavior with LBM, and therefore removed these associations from analyses.

We performed Steiger filtering as a sensitivity analysis to remove genetic variants explaining greater variance in the exposure trait compared to the outcome. To account for likely biases from the *APOE* variant as previously described (22), we performed sensitivity analyses removing IVs within 500kb of the *APOE* rs429358 variant. Briefly, the rs429358 variant is strongly associated with LDL-C and Alzheimer’s disease meaning that participants (especially older ones) with these risk variants and who are still alive and incorporated in the UKB are likely enriched for healthy behaviors that have counter-balanced their genetic risk from *APOE*, leading to a form of survival/selection bias.(22) It is therefore strongly suspected that the association of the *APOE* variant with self-reported MVPA is an artifact of this bias.

### Analyses

MR associations were first estimated with the inverse variance weighted (IVW) linear regression. MVMR analyses were also conducted to address potential confounding of the PA-body composition association by educational attainment. Causal estimates were reported as regression coefficients with 95% confidence intervals (95% CI). Body composition outcomes reflect a one-unit incremental change in self-reported and accelerometer-based exposure variables. Post hoc sensitivity analyses were performed using MR-Egger, median and mode-based estimates, Mendelian randomization pleiotropy residual sum and outlier (MR-PRESSO) (45) and casual analysis using summary effect estimates (CAUSE).(46)

The validity of MR estimates are dependent on three core assumptions: (1) the relevance assumption, meaning selected variants must be directly associated with the exposure; (2) the independence assumption that unmeasured confounders are not associated with the outcome; (3) and the exclusion restriction assumption that genetic variants are not pleiotropically associated with the exposure.

We used the conventional F-statistic threshold of 10 to evaluate instrument strength. We performed visual assessments of IVW regression, funnel plots and leave-one-out (LOO) analyses to identify individual variants influencing causal effects. Heterogeneity was assessed for all MR and MVMR analyses using a modified Cochran’s Q statistic.(47) Outcome and exposure measures were reversed to evaluate for bidirectionality in MR. Associations were considered (statistically) significant at the p<0.05 threshold with no correction for multiple testing. Although we recognize that multiple tests are being performed, we evaluate our results based on the consistency of estimates across multiple exposures, outcomes, and sensitivity tests. MR analyses and quality control procedures were performed using the “TwoSampleMR” (48), “MVMR” (49), and “cause” R packages.(46) Analyses were performed in R v3.6.3 (R Foundation for Statistical Computing).(50)

## RESULTS

GWAS for self-reported moderate PA revealed one novel genome-wide significant SNP, rs6545389 (p = 3.5×10^−8^). GWAS of accelerometer-measured MVPA revealed two novel loci including rs17443704 (p = 3.8×10^−9^) and rs4480415 (p = 8.7×10^−9^), as well as a near genome-wide significant effect for rs791273 (p = 5.0×10^−8^). Additional details on these variants are provided in our supplementary results (Figures S1-2).

Univariable MR analyses suggested self-reported sedentary behavior was positively associated with anthropometric indices and VAT, while no association was identified with LBM (Figure 2). After controlling for educational attainment in MVMR, sedentary behavior associations were null for all measures except an inverse association with WHR (*β =* -0.25, 95%CI: -0.02, -0.48), and a positive association with VAT (*β* = 0.38, 95%CI: 0.27, 0.49) (Figure 3). Self-reported sedentary behavior was significantly associated with VAT across sensitivity analyses including CAUSE (γ = 0.23, 95%CI: 0.19, 0.28) (Table 1). MR analyses for the accelerometer-measured sedentary phenotype similarly identified a significant positive association with BMI (*β* = 0.36, 95%CI: 0.02, 0.71), while other anthropometric outcomes and VAT were null (Figure 4). CAUSE did not identify an association between accelerometer-measured sedentary behavior and VAT (Table 1). After controlling for educational attainment, however, no associations were identified for the accelerometer-measured sedentary phenotype (Figure 5).

**Table 1.** Mendelian Randomization results for the putative effect of sedentary behavior and physical activity on visceral adipose tissue

| Exposure | Self-report |  |  | Accelerometer |  |  |
| --- | --- | --- | --- | --- | --- | --- |
|  | SNPs (#) | Effect | 95% CI | SNPs (#) | Effect | 95% CI |
| **Sedentary Behavior** |  |  |  |  |  |  |
| IVW<sup>††</sup> | 128 | 0.24<sup>***</sup> | 0.20, 0.29 | 4 | 0.15 | -0.01, 0.31 |
| MVMR<sup>††</sup> |  | 0.38<sup>***</sup> | 0.27, 0.49 |  | 0.04 | -0.14, -0.23 |
| MR Egger |  | 0.34<sup>**</sup> | 0.13, 0.56 |  | 0.00 | -0.69, 0.69 |
| Weighted Mode |  | 0.24<sup>***</sup> | 0.13, 0.36 |  | 0.03 | -0.17, 0.24 |
| Weighted Median |  | 0.24<sup>***</sup> | 0.20, 0.28 |  | 0.07 | -0.07, 0.22 |
| MR-PRESSO<sup>§§</sup> |  | 0.30<sup>***</sup> | 0.21, 0.38 |  | 0.12 | -0.06, 0.30 |
| CAUSE<sup>¶</sup> |  | 0.23<sup>***</sup> | 0.19, 0.28 |  | 0.1 | -0.01, 0.21 |
| **Moderate PA** |  |  |  |  |  |  |
| IVW<sup>††</sup> | 32 | 0.08 | -0.01, 0.17 |  | -- |  |
| MVMR<sup>††</sup> |  | 0.14 | -0.07, 0.36 |  | -- |  |
| MR Egger |  | 0.08 | -0.21, 0.38 |  | -- |  |
| Weighted Mode |  | 0.04 | -0.13, 0.21 |  | -- |  |
| Weighted Median |  | 0.03 | -0.05, 0.12 |  | -- |  |
| MR-PRESSO<sup>§§</sup> |  | 0.13 | -0.08, 0.33 |  | -- |  |
| CAUSE<sup>¶</sup> |  | 0.25 | 0.02, 0.47 |  | -- |  |
| **MVPA<sup>†</sup>** |  |  |  |  |  |  |
| IVW<sup>††</sup> | 93 | -0.09<sup>**</sup> | -0.14, -0.03 | 46 | -0.14<sup>***</sup> | -0.19, -0.08 |
| MVMR<sup>††</sup> |  | 0.05 | -0.09, 0.19 |  | -0.12<sup>**</sup> | -0.19, -0.04 |
| MR Egger |  | -0.18 | -0.4, 0.03 |  | -0.09 | -0.24, 0.06 |
| Weighted Mode |  | -0.04 | -0.17, 0.09 |  | -0.03 | -0.18, 0.11 |
| Weighted Median |  | -0.07 | -0.12, -0.01 |  | -0.09<sup>**</sup> | -0.16, -0.03 |
| MR-PRESSO<sup>§§</sup> |  | -0.08 | -0.21, 0.05 |  | 0.00 | -0.03, 0.03 |
| CAUSE<sup>¶</sup> |  | -0.06 | -0.16, 0.04 |  | -0.11<sup>***</sup> | -0.15, -0.07 |
| **Vigorous PA** |  |  |  |  |  |  |
| IVW<sup>††</sup> | 61 | -0.31<sup>***</sup> | -0.31, -0.13 | 26 | -0.19<sup>***</sup> | -0.25, -0.13 |
| MVMR<sup>††</sup> |  | -0.02 | -0.34, 0.29 |  | -0.15<sup>***</sup> | -0.24, -0.07 |
| MR Egger |  | 0.05 | -0.2, 0.3 |  | -0.24 | -0.51, 0.02 |
| Weighted Mode |  | -0.09 | -0.34, 0.15 |  | -0.23 | -0.39, -0.06 |
| Weighted Median |  | -0.14<sup>**</sup> | -0.24, -0.04 |  | -0.19<sup>***</sup> | -0.28, -0.1 |
| MR-PRESSO<sup>§§</sup> |  | -0.12 | -0.40, 0.16 |  | -0.09 | -0.16, -0.02 |
| CAUSE<sup>¶</sup> |  | -0.13 | -0.33, 0.06 |  | -0.14<sup>***</sup> | -0.19, -0.08 |
| **SSOE<sup>‡</sup> / Overall PA<sup>§</sup>** |  |  |  |  |  |  |
| IVW<sup>††</sup> | 98 | -0.37<sup>***</sup> | -0.47, -0.27 | 6 | -0.28 | -0.49, -0.06 |
| MVMR<sup>††</sup> |  | -0.27 | -0.52, -0.01 |  | -0.21 | -0.44, 0.02 |
| MR Egger |  | -0.09 | -0.45, 0.26 |  | -0.25 | -1.08, 0.57 |
| Weighted Mode |  | -0.37 | -0.73, 0.0 |  | -0.17 | -0.39, 0.06 |
| Weighted Median |  | -0.31<sup>***</sup> | -0.41, -0.21 |  | -0.21 | -0.38, -0.03 |
| MR-PRESSO<sup>§§</sup> |  | -0.33<sup>**</sup> | -0.55, -0.11 |  | -0.05 | -0.16, 0.05 |
| CAUSE<sup>¶</sup> |  | -0.28 | -0.46, -0.11 |  | -0.08 | -0.19, 0.03 |

Abbreviations: MVPA, moderate-to-vigorous PA; SSOE, strenuous sports and other exercises; IVW, Inverse Variance Weighted; MVMR, Multivariable MR; MR-PRESSO, pleiotropy residual sum and outlier; CAUSE, causal analysis using summary effect. ‡ Analyses only conducted for self-report PA.
§ Analyses only conducted for accelerometer-measured PA. Note: \*\*\*p<.001, \*\*p<.01

**Figure 2.**
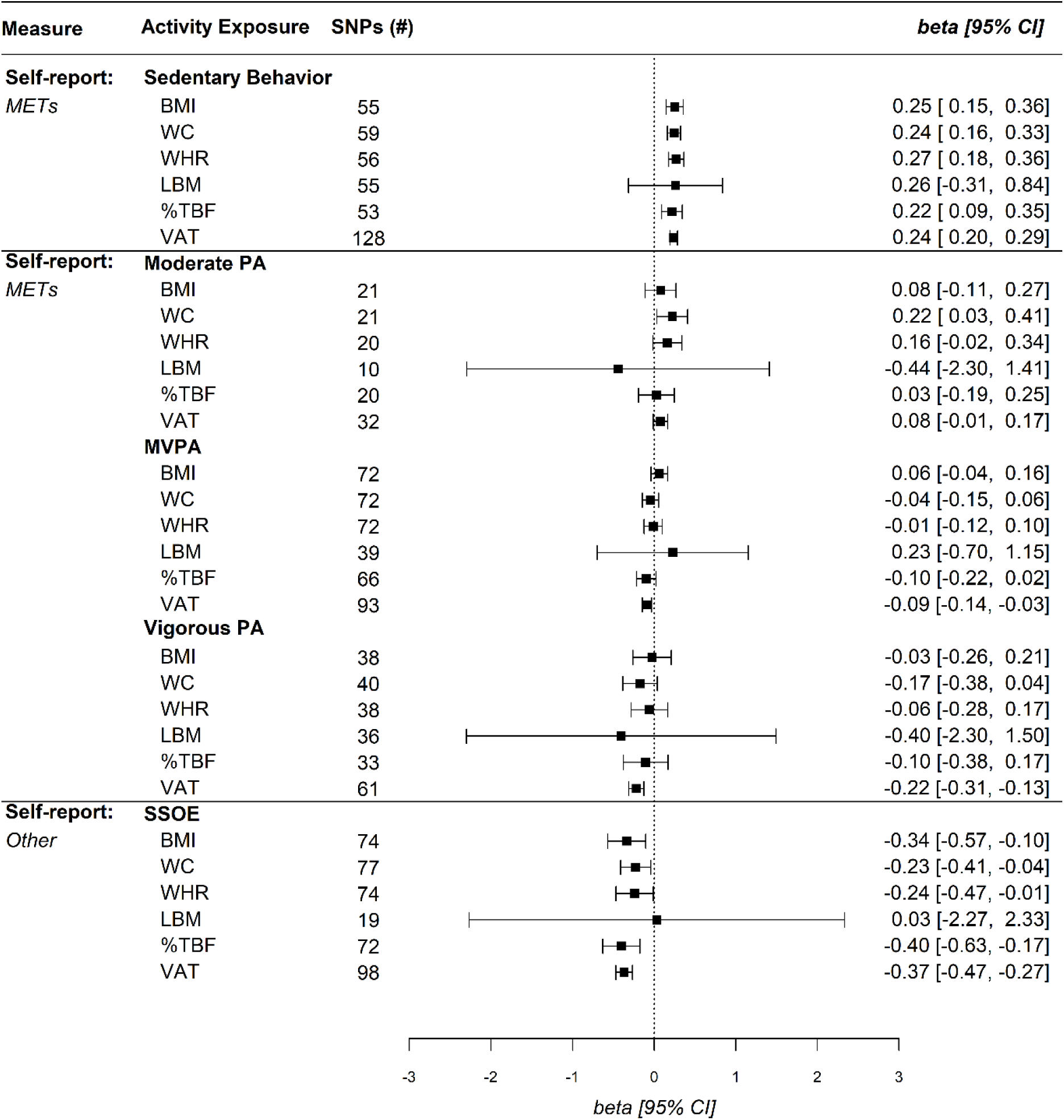
Univariable Mendelian Randomization IVW Estimates for Self-Reported Physical Activity Levels and Body Composition. Abbreviations: SNPs, single nucleotide polymorphisms; CI, confidence interval; METs, metabolic equivalent of tasks; BMI, body mass index; WC, waist circumference; WHR, waist-to-hip ratio; LBM, lean body mass; %TBF, total body fat percent; VAT, visceral adipose tissue; PA, Physical Activity; MVPA, moderate-to-vigorous PA; SSOE, strenuous sports or other exercises

**Figure 3.**
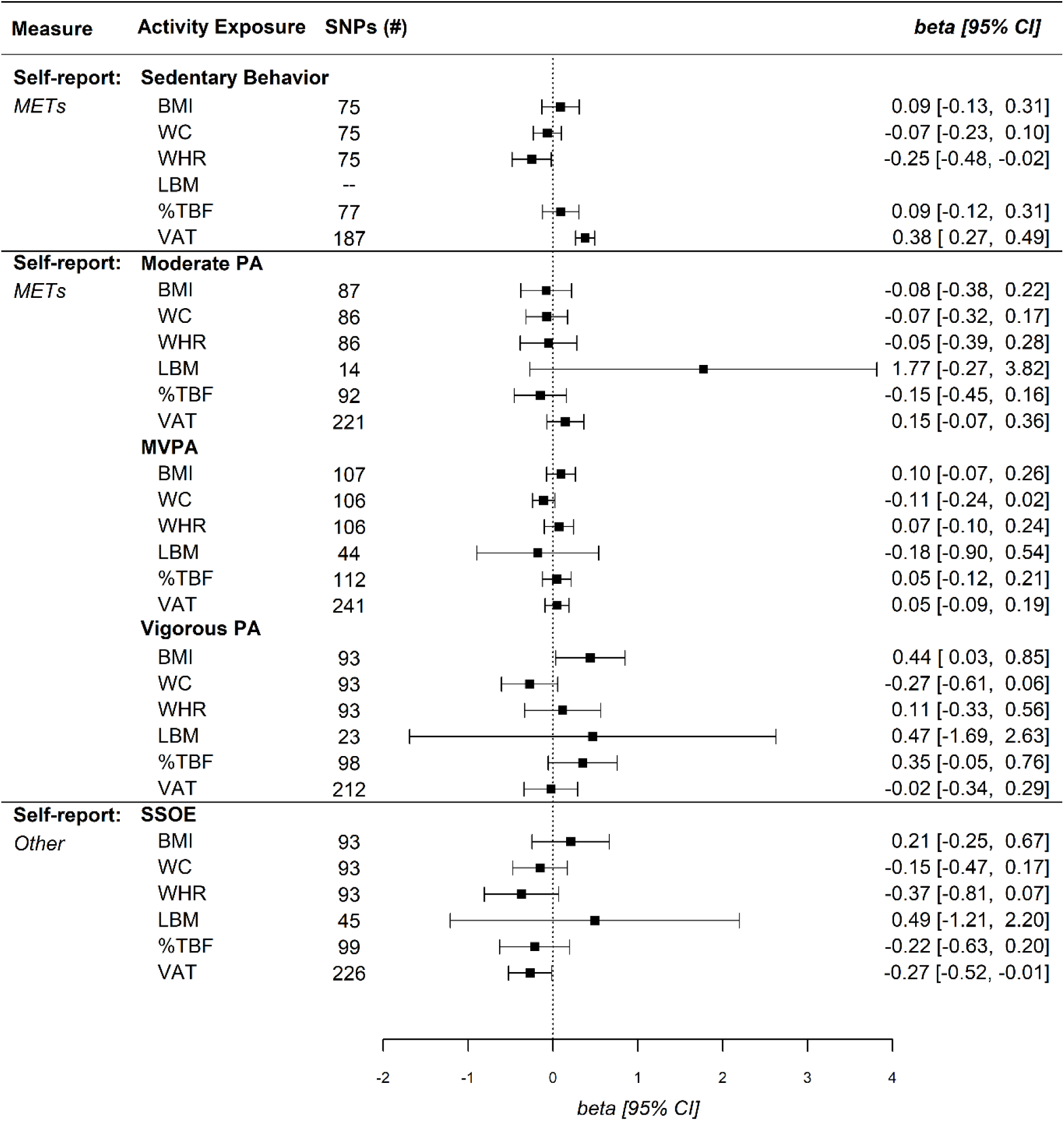
Multivariable Mendelian Randomization IVW Estimates for Self-Reported Physical Activity Levels and Body Composition. Due to an insufficient number of overlapping SNPs for sedentary behavior and LBM with educational attainment, no estimate was identified. Abbreviations: SNPs, single nucleotide polymorphisms; CI, confidence interval; METs, metabolic equivalent of tasks; BMI, body mass index; WC, waist circumference; WHR, waist-to-hip ratio; LBM, lean body mass; %TBF, total body fat percent; VAT, visceral adipose tissue; PA, Physical Activity; MVPA, moderate-to-vigorous PA; SSOE, strenuous sports or other exercises

**Figure 4.**
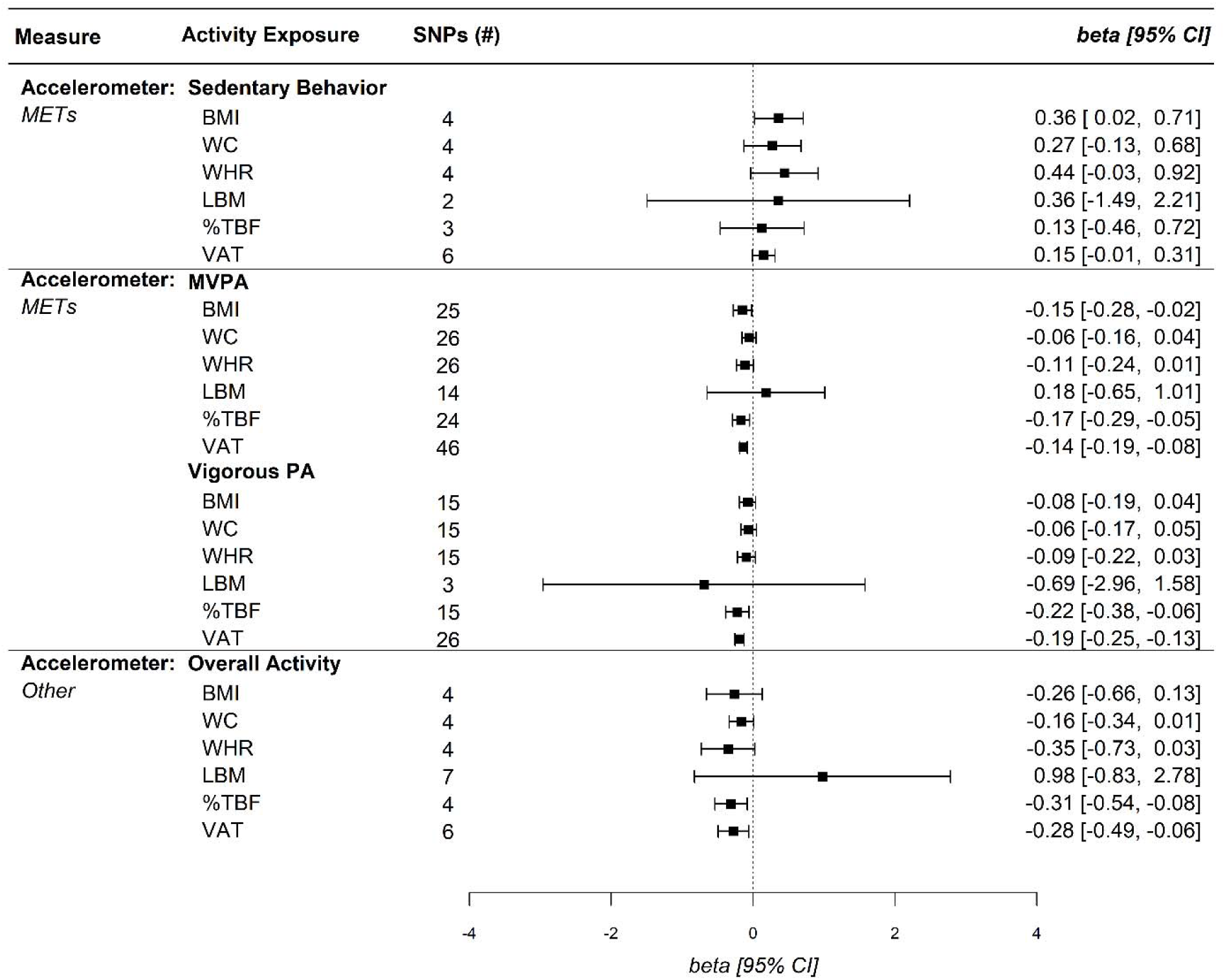
Univariable Mendelian Randomization IVW Estimates for Accelerometer Measured Physical Activity Levels and Body Composition. Abbreviations: SNPs, single nucleotide polymorphisms; CI, confidence interval; METs, metabolic equivalent of tasks; BMI, body mass index; WC, waist circumference; WHR, waist-to-hip ratio; LBM, lean body mass; %TBF, total body fat percent; VAT, visceral adipose tissue; PA, Physical Activity; MVPA, moderate-to-vigorous PA

**Figure 5.**
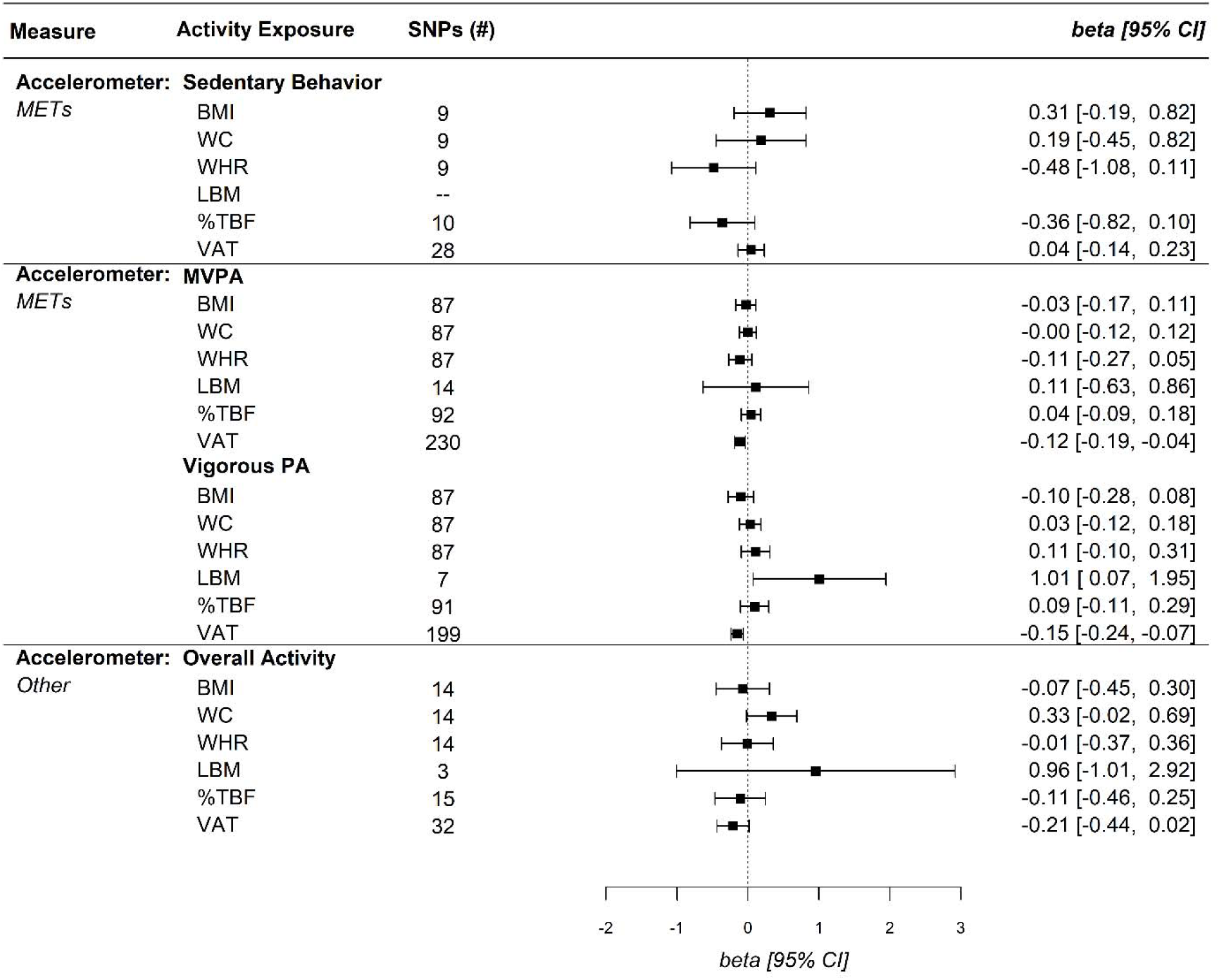
Multivariable Mendelian Randomization IVW Estimates for Accelerometer Measured Physical Activity Levels and Body Composition. Due to an insufficient number of overlapping SNPs for sedentary behavior and LBM with educational attainment, no estimate was identified. Abbreviations: SNPs, single nucleotide polymorphisms; CI, confidence interval; METs, metabolic equivalent of tasks; BMI, body mass index; WC, waist circumference; WHR, waist-to-hip ratio; LBM, lean body mass; %TBF, total body fat percent; VAT, visceral adipose tissue; PA, Physical Activity; MVPA, moderate-to-vigorous PA

MR for self-reported moderate PA revealed a singular positive association with WC (*β = 0*.22, 95%CI: 0.03, 0.41) (Figure 2), while no associations were identified in MVMR. MR for self-reported MVPA was not associated with WC (*β* = -0.10, 95%CI: -0.22, 0.02) while a significant association was identified with VAT (*β* = -0.09, 95%CI: -0.14, -0.03). However, MVMR analyses did not identify a protective association of MVPA on any body composition measure (Figure 3). MR analysis of accelerometer-measured MVPA was negatively associated with %TBF and VAT (Figure 4). After controlling for educational attainment accelerometer-measured MVPA was only protective against VAT (*β* = -0.12, 95%CI: -0.19, -0.04) (Figure 5). We additionally identified a negative association between accelerometer-measured MVPA and VAT in CAUSE (γ = 0.23, 95%CI: 0.19, 0.28) (Table 1).

MR analysis of self-reported vigorous PA suggested a protective association with VAT, while no relationship was identified in MVMR analysis (*β* = -0.02, 95%CI: -0.34, 0.29) (Figure 3). CAUSE did not suggest an association between vigorous PA and VAT (*γ* = -0.13, 95%CI: -0.33, 0.06) (Table 1). In contrast to VAT, no association was identified between vigorous PA and BMI in MR, while MVMR indicated a positive association with BMI (*β* = 0.44, 95%CI: 0.03, 0.85) (Figure 3). No associations were identified between self-reported vigorous PA and other body composition measures. MR analysis of accelerometer measured vigorous PA demonstrated a protective association with %TBF (*β* = -0.22, 95%CI: -0.38, -0.06) and VAT (*β* = -0.19, 95%CI: -0.25, -0.13) (Figure 4). In MVMR analysis, only vigorous PA remained protective against VAT (*β* = -0.15, 95%CI: -0.24, -0.07), while no association was identified for %TBF (*β* = 0.09, 95%CI: -0.11, 0.29) (Figure 5). CAUSE similarly suggested a positive association between accelerometer-measured vigorous PA and VAT (*γ* = -0.14, 95%CI: -0.19, -0.08) (Table 1).

MR analyses with the self-reported SSOE measure demonstrated protective effects for all body composition measures except LBM (Figure 2). In MVMR analyses, all SSOE - body composition associations were null except for VAT (*β* = -0.27, 95%CI: -0.52, -0.01) (Figure 3). CAUSE analyses indicated a negative association between SSOE and VAT (*γ* = -0.28, 95%CI: -0.46, -0.11) (Table 1). MR analyses of the overall activity phenotype (average accelerations) identified significant positive associations with %TBF (*β* = -0.31, 95%CI: -0.54, -0.08) and VAT (*β* = -0.28, 95%CI: -0.49, -0.06) (Figure 4). MVMR and CAUSE did not support a significant protective effect for overall activity on VAT (Table 1).

The F-statistics were greater than 20 for all genetic instruments suggesting instruments were sufficiently associated with the exposure (Table S2). We did not identify genetic variants in visual assessments of regressions, funnel plots, or LOO which may invalidate the independence assumption. The MR-Egger test detected significant pleiotropy for self-reported sedentary behavior and %TBF, self-reported vigorous PA and WHR, and accelerometer-measured vigorous PA and LBM (Table S3). We did not identify other self-reported or accelerometer-measured associations which may have invalidated the exclusion criteria. However, heterogeneity estimates measured by Cochran’s Q indicated widespread pleiotropy across self-reported and accelerometer-measured MR analyses (Table S4). An exception was noted in univariable MR analyses for self-reported PA with %TBF outcome, in which there was little evidence of heterogeneity. In MVMR analyses, there was a notably higher degree of heterogeneity.

MR-PRESSO sensitivity analyses identified some pleiotropic SNPs, which were removed, and evaluated for significance after outlier correction (Table S5). Overall, MR-PRESSO estimates were consistent with our MVMR findings. Self-reported sedentary behavior was positively associated with all body composition outcomes. Although no association was observed for any self-reported vigorous PA exposure, negative associations were identified in accelerometer-measured vigorous PA for %TBF and VAT. Further, we identified robust associations of SSOE with all body composition measures, including VAT.

Other sensitivity analyses including median and mode-based estimates were typically consistent with our main findings (Table S6-7), while MR-Egger estimates trended towards the null (Table S8). CAUSE suggested robust protective effects for accelerometer-measured MVPA and Vigorous PA on VAT, with limited evidence indicating a protective effect of SSOE on VAT (Table S9). CAUSE additionally suggested a positive association of self-reported sedentary behavior on BMI, WHR, WC and VAT, while no effects were identified using the accelerometer-measured sedentary phenotype (Table S9). Reversing our phenotypes did not reveal consistent associations between body composition and PA outcomes (Table S10). Sensitivity analyses excluding the *APOE* rs429358 variant attenuated multiple associations, including the protective effect of accelerometer measured MVPA on VAT. However, accelerometer-measured vigorous PA remained protective for VAT (Figure S1-2), and lower intensity levels of PA continued to show no association with anthropometric or body composition measures. We did not identify directional changes for any associations after Steiger filtering sensitivity analyses. Across self-report and accelerometer PA exposures sensitivity analyses, including CAUSE and MVMR, our results suggest an association between genetically-predicted higher-intensity PA with lower VAT (Table 1).

## DISCUSSION

Ambiguity currently exists in our understanding of whether PA protects against obesity and/or central adiposity, which has been driven by fundamental limitations to both observational studies and intervention trials.(8,9,29) The relationship between PA and body composition traits is highly complex, and genetic variants underlying each of these have been shown to have pleiotropic effects, including with each other (e.g. *CADM2* associated with both BMI and PA). Therefore, efforts are needed to better elucidate the shared genetics and pathways that link PA with weight and body composition.(51) We applied MR using a comprehensive set of PA and body composition measures to increase confidence in causal estimations between PA and body composition, considering self-reported and accelerometer-based PA exposures in parallel analyses. Our results provide some evidence of a protective effect on VAT from genetically-predicted levels of accelerometer-measured MVPA and vigorous PA, as well as self-reported SSOE. In contrast to these findings with VAT, we find limited evidence of a causal relationship between genetically-predicted PA and other body composition indexes.

MR estimates were not always consistent across self-reported and accelerometer-based PA measures. Self-reported measures may not capture some aspects of activity or sedentary behavior, while accelerometer measurements were designed to record uninterrupted activity.(52) Further, accelerometer measurements reflected activity from one specific week and may not capture some forms of PA such as resistance training.(37) Overall, each of these methods likely suffers from both shared and unique biases. These two types of measures can thus be thought of as complementary, and evidence supported by both measures may be viewed as stronger.

MR and MVMR analyses suggested consistent protective associations for accelerometer-measured MVPA, vigorous PA, and self-reported SSOE with VAT. Sensitivity analyses with MR-PRESSO, median, and mode-based estimates were consistent with the direction of IVW estimates despite the reduced sensitivity of these methods. CAUSE estimates were marginally attenuated compared to other methods, but similarly suggested a protective association from higher-intensity PA. Although we detected a strong positive association between self-reported sedentary behavior and VAT, acceleration-measured sedentary exposure did not reveal any association with VAT. Although the directional effect was stable between self-report and accelerometer measurements, we find limited evidence suggesting a consistent positive association between sedentary behavior and VAT. Further, we did not observe any consistent protective associations between moderate PA with VAT. Overall, these associations suggest an independent effect of high-intensity PA on VAT.

Our results align with previous evidence suggesting an independent protective effect of PA on VAT. The over-proportional reduction of VAT compared to subcutaneous tissues may be explained by heterogeneous lipolysis regulation, which has been shown to be dependent on the location of adipose tissue in the body.(53) Increasing levels of PA intensity and duration has been shown to increase secretion of catecholamines.(54) In contrast to subcutaneous tissue types, higher concentrations of *β*- and α2A-adrenoceptors in VAT enhances lipolysis through binding of hormones including catecholamines, thereby upregulating cyclic AMP production and increasing hydrolysis of triglycerides.(55) Other physiological features such as reduced insulin receptor substrate-1 in VAT limits antilipolytic processes by lowering insulin sensitivity in this region compared to subcutaneous tissue.(55) Increased blood flow and higher concentration of cells in the abdominal cavity may further amplify regional lipolytic activity. In a previous intervention by Johnson et al. (56) aerobic exercise was shown to reduce visceral adiposity without change to BMI or %TBF. A systematic review of randomized PA control trials further identified a small number of studies where VAT was reduced independently from body mass changes.(57) Here, we find evidence suggesting that PA at sufficient levels of intensity may independently reduce VAT and improve an individual’s metabolic profile.

We did not identify consistent associations between genetically-predicted PA exposures and other body composition outcomes. Although MR analyses indicated positive associations of sedentary behavior with multiple body composition measures, no positive associations were identified for body composition measures (with the exception of VAT) after controlling for educational attainment. Similarly, MR estimates of accelerometer-measured sedentary behavior were null. MR estimates of higher-intensity MET-based measures, including MVPA and vigorous PA, revealed few protective effects for anthropometric measures. While both accelerometer-measured MVPA and vigorous PA demonstrated negative associations with %TBF, these effects were again null after controlling for educational attainment. MVMR estimates for exposures not based on METs, including SSOE and overall activity, did not reach significance. Sensitivity analyses using MR-PRESSO, median and mode-based estimates were similarly inconsistent or null across sedentary behavior, moderate PA, MVPA, and vigorous PA exposures. In contrast to other null body composition findings, we identified a positive association with genetically-predicted self-reported vigorous PA and BMI, and accelerometer-measured PA and LBM in MVMR analysis. These results may suggest that PA has a direct positive effect on BMI and LBM. We interpret the robustness of these findings with caution, however, due to the null effects in parallel analyses and other sensitivity tests.

Our findings contrast with bidirectional two-sample MR analyses from Carrasquilla et al. (58) who used summary statistics identical to our study including self-reported vigorous PA, accelerometer-measured moderate PA and BMI. While Carrasquilla et al. reported significant IVW and CAUSE associations for moderate and vigorous PA on BMI, we did not identify a similar effect in IVW, MVMR or CAUSE analyses. These discrepancies may have been a result of the differing instrument selection thresholds and the modification of priors used in CAUSE between studies. A previous MR analysis from Doherty et al. (23) additionally identified evidence suggesting an inverse bidirectional relationship between BMI and accelerometer-measured overall activity. After controlling for educational attainment, however, we did not identify a negative association between PA and BMI. Further, bidirectional IVW estimates in our study did not suggest a causal association of BMI, %TBF or LBM on PA. In contrast to our study, Doherty and colleagues used an accelerometer-measured overall activity exposure distinct from our own exposure, as well as a likelihood-based method to estimate causal effects. A MR meta-analysis from Shnurr et al. (51) using a smaller cohort of both children and adults further suggested a positive association of BMI on sedentary time. We identified a consistent positive association for BMI on self-reported sedentary behavior, while accelerometer-measured estimates of BMI on sedentary behavior were null. Similar to Shnurr et al. (51), we did not identify an effect of genetically-predicted BMI on overall activity or MVPA. Overall, our findings conflicting with previous literature may have been driven by differing instrumental variable inclusion criteria, as was well as application of more conservative methodologies such as MVMR and CAUSE.

Health agencies uniformly emphasize the importance of moderate PA for weight management.(59,60) Some evidence suggests, however, that these PA recommendations are insufficient.(61) While there is little doubt that PA offers immensurable benefits to overall health, our results suggest a limited protective effect for anthropometric indices, LBM, or %TBF from PA. Our null findings may be plausible under the recently proposed constrained total energy expenditure (TEE) model.(62) In contrast to conventional additive models, the constrained TEE model accounts for potential downregulation of non-musculoskeletal physiological activity during higher levels of PA, resulting in TEE within a homeostatically constrained range.(63) This model may provide an explanatory framework for the lack of associations observed between PA and most body composition measures in our analyses.

### Limitations

Our study includes several critical limitations. First, we were unable to identify SNPs at the genome-wide significance level in GWAS and used a reduced instrumental variable threshold (p < 5×10^−6^) across our analyses. The lower predictive capacity of these SNPs may have limited our ability to detect significant sedentary and PA effects on body composition. Despite this limitation, all IVs indicated sufficient strength, as measured by the F-statistic. Additionally, while we applied two-sample MR to limit weak instrument bias, stratification of VAT into samples not overlapping with sedentary behavior or PA may have introduced a form of selection bias. The inherent assumptions of MR may be violated when evaluating polygenic risk factors, such as PA, as the influence of genetic variants on such exposures is unlikely to be specific. We therefore applied MVMR to estimate the direct causal effect of sedentary and PA exposures, which is valuable when instruments are suspected to violate the second and third instrumental variable assumptions.(47) The lower number of instruments identified for accelerometer-measured exposures may have reduced the ability to detect significant associations in MVMR; however, we note that null findings were consistent with CAUSE analyses.

A key design limitation was our inability to evaluate light PA exposures in self-reported or accelerometer-based measurements. Despite the large consortia used in this investigation, we were unable to perform sex-stratified analysis due to limited instrument availability. Our results may have been further attenuated by effect modification, as both regional adiposity (64) and PA (38) have been shown to differ between sexes. We acknowledge a potential increase in the Type I error rate in analyses on account of no correction for multiple testing. Here we report and consider each association individually.(65) Importantly, we do not rely on a single estimate for our inferences, but more broadly consider the consistency of associations across measures and methods to draw conclusions. We emphasize that MR does not provide an infallible test of causation and encourage replication in future studies.

In conclusion, MR and MVMR analyses of various PA levels did not demonstrate consistent protective effects on body composition measures. Our null findings reaffirm the equivocal effectiveness of weight management programs relying on PA alone, in contrast to programs utilizing both dietary restriction and PA. We do find evidence of an association between genetically-predicted high-intensity PA and lower VAT. At the time of this work, no peer-reviewed predictive models for subcutaneous adipose tissue, ectopic fat, or hepatic fat were identified to evaluate as an outcome with PA. MR analyses of these and other more detailed body composition outcomes may provide greater insight into specific cardiometabolic benefits of PA.

## Data Availability

GWAS summary statistics derived as part of this study will be made publicly available in the GWAS Catalog.

## ACKNOWLEDGEMENTS

We would like to recognize the UK Biobank, and the GIANT and GEFOS consortia for their incredible efforts in developing and curating the data sources used in this study.

## DATA SHARING

All code used to conduct analyses and generate figures can be found at https://osf.io/uc4jz/. GWAS summary statistics derived as part of this study will be made publicly available in the GWAS Catalog.

