## supplemental materials for "Association of Sedentary and Physical Activity Behaviors with Body Composition: a Genome-Wide Association and Mendelian Randomization Study"

### Supplemental Tables and Figures

**Figure S3.** Manhattan Plot and genome-wide significant SNPs for self-reported moderate PA ................ **Error! Bookmark not defined.**

**Figure S4.** Manhattan Plot and genome-wide significant SNPs for accelerometer-measured MVPA ........ **Error! Bookmark not defined.**

**Table S1.** Participant Characteristics Included in the Genome-Wide Association Studies

| Phenotype | Study | n | Women (%) | Age, Years (mean ± SD) | Trait Value (mean ± SD) |
|---|---|---|---|---|---|
| **Self-report (hr./day)** | | | | | |
| Sedentary Behavior | UKB / van de Vegte et al. | 422,218 | 54.3 | 57.4 ± 8.0 | 4.7 ± 2.7 |
| Moderate PA | UKB / Novel | 360,912 | 53.4 | 56.7 ± 8.1 | 1.2 ± 1.3 |
| MVPA | UKB / Klimentidis et al. | 377,234 | 50.6 | 56.1 ± 8.2 | 1.9 ± 1.8 |
| Vigorous PA (*≥3 vs. 0 dys./wk.*)† | UKB / Klimentidis et al. | 98,060; 162,995 | 47.3; 58.1 | 55.4 ± 8.3; 57.4 ± 7.8 | 1.1 ± 0.9; 0.0 ± 0.0 |
| SSOE (*>2-3 vs 0 dys./wk.*)† | UKB / Klimentidis et al. | 124,842; 225,650 | 38.1; 35.3 | 52.7 ± 8.1; 52.6 ± 7.9 | 1.1 ± 0.7; 0.0 ± 0.0 |
| **Accelerometer** | | | | | |
| Sedentary Behavior§ | UKB / Doherty et al. | 91,105 | -- | -- | -- |
| MVPA‡ | UKB / Novel | 97,737 | 56.1 | 56.2 ± 7.8 | 0.0799 ± 0.0384 |
| Vigorous PA‡ | UKB / Klimentidis et al. | 90,667 | 56.1 | 56.2 ± 7.8 | 0.0026 ± 0.0033 |
| Overall activity (*milli-gravities*) | UKB / Klimentidis et al. | 91,105 | 56.1 | 56.2 ± 7.8 | 27.1 ± 27.0 |
| **Body Comp.** | | | | | |
| BMI (*kg/m²*) †† | GIANT / Locke at al. | 249,796 | -- | -- | -- |
| WHR†† | GIANT/ Shungin at al. | 224,459 | -- | -- | -- |
| WC (*cm*)†† | GIANT/ Shungin at al. | 224,459 | -- | -- | -- |
| LBM (*kg*) †† | GEFOS / Zillikens et al. | 38,292 | -- | -- | -- |
| %TBF | Lu et al. | 89,297 | 48.3 | 57.8 ± 9.4 | 31.6 ± 6.7 |
| VAT (Self-Report PA; *kg*) | UKB / Karlsson et al. | 56,908 | 56.7 | 57.1 ± 7.8 | 1.5 ± .86 |
| VAT (Accel. PA; *kg*) | UKB / Karlsson et al. | 323,769 | 53.8 | 56.9 ± 8.1 | 1.4 ± .83 |
| **Covariate** | | | | | |
| Educational Attainment | UKB / Okbay et al. | 293,723 | 55.1 | 57.5 ± 8.0 | 14.3 ± 3.6 |

Abbreviations: PA, Physical Activity, MVPA, moderate-to-vigorous PA; SSOE, strenuous sports or other exercises; BMI, body mass index; WC, waist circumference; WHR, waist-to-hip ratio; LBM, lean body mass; %TBF, total body fat percent; VAT, visceral adipose tissue; UKB, UK Biobank, GIANT, Genetic Investigation of ANthropometric Traits consortium; GEFOS, GEnetic Factors for Osteoporosis consortium.

†Phenotype was evaluated as a binary variable.

§Phenotype was trained on 153 free-living individuals to predict metabolic equivalent of tasks.

‡Phenotype was derived from fraction accelerations based on metabolic equivalent of tasks.

††Consortium demographics unavailable.

**Table S2.** F Statistics for Mendelian Randomization Analyses for Physical Activity to Body Composition

| **Phenotypes** | | **Self-report MR** | | | **Accelerometer MR** | | |
|---|---|---|---|---|---|---|---|
| *Activity* | *Body Comp.* | *Mean F* | *Median F* | *Range F* | *Mean F* | *Median F* | *Range F* |
| Sedentary Behavior | LBM | 45.75 | 38.97 | 30.41, 144.19 | 27.61 | 27.61 | 27.26, 27.95 |
| | BMI | 45.32 | 38.55 | 30.41-144.19 | 25.34 | 25.2 | 23.01, 27.95 |
| | WHR | 45.3 | 38.48 | 30.41-144.19 | 25.34 | 25.2 | 23.01, 27.95 |
| | WC | 45.23 | 38.62 | 30.41-144.19 | 25.34 | 25.2 | 23.01, 27.95 |
| | %TBF | 45.77 | 38.97 | 30.41-144.19 | 26.07 | 27.26 | 23.01, 27.95 |
| | VAT | 42.71 | 37.3 | 30.41-144.19 | 26.4 | 26.69 | 23.09, 30.18 |
| Moderate PA | LBM | 23.40 | 23.39 | 21.52, 26.05 | -- | -- | -- |
| | BMI | 23.41 | 23.37 | 21.12-30.38 | -- | -- | -- |
| | WHR | 23.39 | 23.13 | 21.12-30.38 | -- | -- | -- |
| | WC | 23.39 | 23.13 | 21.12-30.38 | -- | -- | -- |
| | %TBF | 23.51 | 23.37 | 21.12-30.38 | -- | -- | -- |
| | VAT | 23.09 | 22.77 | 21.02-30.38 | -- | -- | -- |
| MVPA | LBM | 25.85 | 24.16 | 20.97, 45.99 | 25.13 | 23.32 | 21.62, 33.12 |
| | BMI | 25.94 | 24.41 | 20.88-45.99 | 24.83 | 23.4 | 20.91, 34.67 |
| | WHR | 25.94 | 24.41 | 20.88-45.99 | 24.83 | 23.4 | 20.91, 34.67 |
| | WC | 25.9 | 24.39 | 20.88-45.99 | 24.83 | 23.4 | 20.91, 34.67 |
| | %TBF | 26.02 | 24.39 | 20.88-45.99 | 24.68 | 23.4 | 20.91, 34.67 |
| | VAT | 25.75 | 23.9 | 20.86-45.99 | 24.06 | 23.24 | 20.92, 34.72 |
| Vigorous PA | LBM | 27.78 | 23.05 | 21.18, 55.26 | 23.56 | 22.58 | 20.88, 26.64 |
| | BMI | 26.6 | 23.48 | 20.98-55.26 | 24.98 | 24.56 | 20.88, 36.07 |
| | WHR | 26.73 | 23.54 | 20.98-55.26 | 24.98 | 24.56 | 20.88, 36.07 |
| | WC | 26.47 | 23.42 | 20.98-55.26 | 24.98 | 24.56 | 20.88, 36.07 |
| | %TBF | 27.07 | 23.54 | 20.98-55.26 | 24.98 | 24.56 | 20.88, 36.07 |
| | VAT | 25.62 | 23.05 | 20.9-55.26 | 24.51 | 23.03 | 20.88, 36.76 |
| SSOE | LBM | 25.40 | 24.48 | 20.98, 37.34 | -- | -- | -- |
| | BMI | 26.03 | 24.52 | 20.98-75.4 | -- | -- | -- |
| | WHR | 25.99 | 24.65 | 20.98-75.4 | -- | -- | -- |
| | WC | 26.09 | 24.59 | 20.98-75.4 | -- | -- | -- |
| | %TBF | 26.17 | 24.65 | 20.98-75.4 | -- | -- | -- |
| | VAT | 25.94 | 24.48 | 20.89-82.57 | -- | -- | -- |
| Overall Activity | LBM | -- | -- | -- | 25.16 | 24.84 | 22.66, 27.98 |
| | BMI | -- | -- | -- | 24.54 | 23.76 | 22.66, 27.98 |
| | WHR | -- | -- | -- | 24.54 | 23.76 | 22.66, 27.98 |
| | WC | -- | -- | -- | 24.54 | 23.76 | 22.66, 27.98 |
| | %TBF | -- | -- | -- | 24.54 | 23.76 | 22.66, 27.98 |
| | VAT | -- | -- | -- | 24.45 | 24.84 | 21.28, 27.98 |

Abbreviations: MR, Mendelian randomization; PA, physical activity; MVPA, moderate-to-vigorous PA; SSOE, strenuous sports or other exercises; LBM, lean body mass; BMI, body mass index; WHR, waist-to-hip ratio; WC, waist circumference; %TBF, total body fat percent; VAT, visceral adipose tissue.

Note: An F statistic of 10 indicates that the bias of the IV estimator is 10% of the bias of the observational estimator.

**Table S3.** MR-EGGER Intercept Estimates for Physical Activity to Body Composition

| Phenotypes | | Self-Report MR | | Accelerometer MR | |
|---|---|---|---|---|---|
| *Activity* | *Body Comp.* | *MR-EGGER Intercept* | *p* | *MR-EGGER Intercept* | *p* |
| Sedentary Behavior | LBM | 0.025031 | 0.2958 | -- | -- |
| | BMI | -0.00449 | 0.3034 | 0.017358 | 0.64 |
| | WHR | -0.00209 | 0.5284 | 0.00348 | 0.94 |
| | WC | -0.00449 | 0.2279 | 0.002724 | 0.96 |
| | %TBF | -0.01355 | 0.0073 | 0.052934 | 0.39 |
| | VAT | -0.00168 | 0.586 | 0.005832 | 0.59 |
| Moderate PA | LBM | 0.037154 | 0.5654 | -- | -- |
| | BMI | 0.006266 | 0.3227 | -- | -- |
| | WHR | 0.001847 | 0.7752 | -- | -- |
| | WC | 0.0043 | 0.4634 | -- | -- |
| | %TBF | -0.00217 | 0.7637 | -- | -- |
| | VAT | -0.00245 | 0.6182 | -- | -- |
| MVPA | LBM | 0.035138 | 0.34 | 0.035717 | 0.513 |
| | BMI | 0.001909 | 0.5612 | -0.00662 | 0.36 |
| | WHR | -0.00197 | 0.5577 | -0.00404 | 0.47 |
| | WC | 0.002466 | 0.496 | -0.00533 | 0.44 |
| | %TBF | 0.000303 | 0.9364 | -0.00891 | 0.18 |
| | VAT | 0.004179 | 0.2116 | -0.00181 | 0.39 |
| Vigorous PA | LBM | 0.013209 | 0.753 | -0.41894 | 0.047 |
| | BMI | -0.0054 | 0.216 | 0.004192 | 0.74 |
| | WHR | -0.00829 | 0.0331 | 0.004787 | 0.7 |
| | WC | -0.0051 | 0.2259 | -0.00019 | 0.99 |
| | %TBF | -0.00134 | 0.7813 | 0.026992 | 0.13 |
| | VAT | -0.00144 | 0.7043 | -0.00095 | 0.8 |
| SSOE | LBM | -0.00619 | 0.8902 | -- | -- |
| | BMI | -0.00428 | 0.3557 | -- | -- |
| | WHR | -0.00214 | 0.549 | -- | -- |
| | WC | -0.00288 | 0.5143 | -- | -- |
| | %TBF | -0.00267 | 0.5652 | -- | -- |
| | VAT | -0.00182 | 0.5187 | -- | -- |
| Overall Activity | LBM | -- | -- | -0.3933 | 0.631 |
| | BMI | -- | -- | -0.1215 | 0.32 |
| | WHR | -- | -- | -0.06486 | 0.29 |
| | WC | -- | -- | -0.13557 | 0.23 |
| | %TBF | -- | -- | -0.03473 | 0.64 |
| | VAT | -- | -- | -0.00733 | 0.28 |

Abbreviations: MR, Mendelian randomization; PA, physical activity; MVPA, moderate-to-vigorous PA; SSOE, strenuous sports or other exercises; LBM, lean body mass; BMI, body mass index; WHR, waist-to-hip ratio; WC, waist circumference; %TBF, total body fat percent; VAT, visceral adipose tissue.

**Table S4.** Heterogeneity estimates Mendelian Randomization Analyses for Physical Activity to Body Composition

| **Phenotypes** | | **Self-report MR** | | **Self-report MVMR** | | **Accelerometer MR** | | **Accelerometer MVMR** | |
|---|---|---|---|---|---|---|---|---|---|
| *Activity* | *Body Comp.* | *Q Stat.* | *p* | *Q Stat.* | *p* | *Q Stat.* | *p* | *Q Stat.* | *p* |
| **Sedentary Behavior** | LBM | 46.9 | 0.7435 | -- | -- | 22.2 | 0.035 | -- | -- |
| | BMI | 139.5 | <0.001 | -- | -- | 18.9 | <0.001 | 16.3 | 0.0121 |
| | WHR | 82.4 | 0.0098 | 253.7 | <0.001 | 27 | <0.001 | 17.5 | 0.0077 |
| | WC | 74 | 0.0766 | 145.1 | <0.001 | 19.9 | <0.001 | 18.2 | 0.0058 |
| | %TBF | 86.3 | 0.002 | 141.1 | <0.001 | 11.8 | 0.0027 | 5.8 | 0.5654 |
| | VAT | 260.6 | <0.001 | 315.8 | <0.001 | 33.1 | <0.001 | 160.8 | <0.001 |
| **Moderate PA** | LBM | 4.2 | 0.8959 | 21.3 | 0.03 | -- | -- | -- | -- |
| | BMI | 33.2 | 0.0321 | -- | -- | -- | -- | -- | -- |
| | WHR | 18.4 | 0.4927 | 279.6 | <0.001 | -- | -- | -- | -- |
| | WC | 24.8 | 0.1661 | 166.1 | <0.001 | -- | -- | -- | -- |
| | %TBF | 13.9 | 0.788 | 150.8 | <0.001 | -- | -- | -- | -- |
| | VAT | 52 | 0.0077 | 477.9 | <0.001 | -- | -- | -- | -- |
| **MVPA** | LBM | 4.2 | 0.8959 | 21.3 | 0.03 | 18.9 | <0.001 | 13.1 | 0.2867 |
| | BMI | 110.8 | <0.001 | -- | -- | 96.9 | <0.001 | -- | -- |
| | WHR | 91.2 | 0.0318 | 286.4 | <0.001 | 66.6 | <0.001 | 271.7 | <0.001 |
| | WC | 94 | 0.0245 | 187 | <0.001 | 44.8 | 0.0089 | 171.8 | <0.001 |
| | %TBF | 59.1 | 0.6155 | 160.6 | <0.001 | 34.3 | 0.0613 | 144.4 | <0.001 |
| | VAT | 202.9 | <0.001 | 554.2 | <0.001 | 183.5 | <0.001 | 1709.7 | <0.001 |
| **Vigorous PA** | LBM | 21.7 | 0.2472 | 37.5 | 0.01 | 13.4 | 0.037 | 1.3 | 0.8616 |
| | BMI | 71.4 | <0.001 | -- | -- | 20.4 | 0.119 | -- | -- |
| | WHR | 47.1 | 0.1242 | 279.8 | <0.001 | 17.8 | 0.2154 | 278.4 | <0.001 |
| | WC | 47.4 | 0.1685 | 153.3 | <0.001 | 13.7 | 0.4738 | 165.1 | <0.001 |
| | %TBF | 34.1 | 0.3687 | 165.8 | <0.001 | 19 | 0.1666 | 162.6 | <0.001 |
| | VAT | 148.8 | <0.001 | 538.1 | <0.001 | 90.8 | <0.001 | 1308.6 | <0.001 |
| SSOE | LBM | 25.5 | 0.8799 | 42.4 | 0.453 | -- | -- | -- | -- |
| | BMI | 190.7 | <0.001 | -- | -- | -- | -- | -- | -- |
| | WHR | 132.2 | <0.001 | 306.1 | <0.001 | -- | -- | -- | -- |
| | WC | 98 | 0.0386 | 173.4 | <0.001 | -- | -- | -- | -- |
| | %TBF | 51.1 | 0.9472 | 188.5 | <0.001 | -- | -- | -- | -- |
| | VAT | 182.3 | <0.001 | 458.4 | <0.001 | -- | -- | -- | -- |
| Overall Activity | LBM | -- | -- | -- | -- | 4.1 | 0.1314 | 1.3 | 0.8616 |
| | BMI | -- | -- | -- | -- | 20.6 | <0.001 | 38 | <0.001 |
| | WHR | -- | -- | -- | -- | 14 | 0.0029 | 35.4 | <0.001 |
| | WC | -- | -- | -- | -- | 2.6 | 0.4596 | 27 | 0.0046 |
| | %TBF | -- | -- | -- | -- | 2.5 | 0.4674 | 15.8 | 0.1986 |
| | VAT | -- | -- | -- | -- | 11.5 | 0.0211 | 214.7 | <0.001 |

Abbreviations: MR, Mendelian randomization; MVMR, Multivariable Mendelian randomization; PA, physical activity; MVPA, moderate-to-vigorous PA; SSOE, strenuous sports or other exercises; LBM, lean body mass; BMI, body mass index; WHR, waist-to-hip ratio; WC, waist circumference; %TBF, total body fat percent; VAT, visceral adipose tissue.

Note: A large Q statistic and small p-value indicates instrument heterogeneity and potential violation of the MR assumptions. Where horizontal pleiotropy does not exist, observed values are expected to have smaller residuals, meaning that the variants do not deviate from the slope of the fitted regression. Inversely, variants deviating from the regression line suggest a pleiotropic effect.

**Table S5.** MR-PRESSO Estimates for Physical Activity to Body Composition

| **Phenotypes** | | **Self-Report MR** | | **Accelerometer MR** | |
|---|---|---|---|---|---|
| *Activity* | *Body Comp.* | *Causal Estimate* | *p* | *Causal Estimate* | *p* |
| Sedentary Behavior | LBM | 0.26 | 0.3437 | 0.19* | 0.0646 |
| | BMI | 0.23 | <0.001 | 0.19* | 0.1228 |
| | WHR | 0.23 | <0.001 | 0.46** | 0.0068 |
| | WC | 0.24 | <0.001 | 0.43* | 0.1844 |
| | %TBF | 0.22 | 0.0011 | 0.02 | 0.8574 |
| | VAT | 0.3 | <0.001 | 0.12* | 0.1826 |
| Moderate PA | LBM | -0.44 | 0.5124 | -- | -- |
| | BMI | 0.08 | 0.4447 | -- | -- |
| | WHR | 0.22 | 0.0426 | -- | -- |
| | WC | 0.16 | 0.09 | -- | -- |
| | %TBF | 0.01 | 0.8836 | -- | -- |
| | VAT | 0.13 | 0.2193 | -- | -- |
| MVPA | LBM | 0.23 | 0.5223 | 0.18 | 0.6794 |
| | BMI | 0.08 | 0.1298 | -0.11* | 0.014 |
| | WHR | -0.05 | 0.3468 | -0.06 | 0.2577 |
| | WC | 0 | 0.9948 | -0.06* | 0.2203 |
| | %TBF | -0.11 | 0.0588 | -0.17 | 0.0114 |
| | VAT | -0.08 | 0.2327 | 0* | 0.9618 |
| Vigorous PA | LBM | 0.03 | 0.9778 | 0.98 | 0.3291 |
| | BMI | -0.11 | 0.3991 | -0.08 | 0.1982 |
| | WHR | -0.23 | 0.0379 | -0.06 | 0.2683 |
| | WC | -0.16 | 0.2133 | -0.09 | 0.1648 |
| | %TBF | -0.1 | 0.4995 | -0.22 | 0.0195 |
| | VAT | -0.12 | 0.3949 | -0.09* | 0.0106 |
| SSOE | LBM | -0.4 | 0.6289 | -- | -- |
| | BMI | -0.49* | <0.001 | -- | -- |
| | WHR | -0.21 | 0.0261 | -- | -- |
| | WC | -0.3* | 0.0124 | -- | -- |
| | %TBF | -0.41 | <0.001 | -- | -- |
| | VAT | -0.33** | 0.003 | -- | -- |
| Overall Activity | LBM | -- | -- | 0.05 | 0.205 |
| | BMI | -- | -- | -0.31 | 0.0638 |
| | WHR | -- | -- | -0.16* | 0.4411 |
| | WC | -- | -- | -0.16 | 0.139 |
| | %TBF | -- | -- | -0.28* | 0.3675 |
| | VAT | -- | -- | -0.05* | 0.3327 |

Abbreviations: MR, Mendelian randomization; PA, physical activity; MVPA, moderate-to-vigorous PA; SSOE, strenuous sports or other exercises; LBM, lean body mass; BMI, body mass index; WHR, waist-to-hip ratio; WC, waist circumference; %TBF, total body fat percent; VAT, visceral adipose tissue.

Note: MR-PRESSO evaluates for global pleiotropic effects by comparing the residual sum of squares with the expected distance under the null hypothesis for no horizontal pleiotropy. Individual pleiotropic outliers are identified by examining the observed and expected distributions of each variant; *Value was corrected for outliers; ** MR-PRESSO estimate reached significance after outlier correction.

**Table S6.** MR Median-based Estimates for Physical Activity to Body Composition

| Phenotypes | | Self-Report MR | | | Accelerometer MR | | |
|---|---|---|---|---|---|---|---|
| *Activity* | *Body Comp.* | *SNPs (#)* | *beta* | *95% CI* | *SNPs (#)* | *beta* | *95% CI* |
| Sedentary Behavior | LBM | 55 | 0.32 | -0.54, 1.18 | 4 | 0.26 | 0.05, 0.46 |
| | BMI | 55 | 0.22 | 0.11, 0.33 | 4 | 0.35 | 0.07, 0.64 |
| | WHR | 56 | 0.24 | 0.12, 0.37 | 4 | 0.42 | 0.21, 0.63 |
| | WC | 59 | 0.22 | 0.11, 0.33 | 3 | -0.13 | -0.5, 0.24 |
| | %TBF | 53 | 0.34 | 0.18, 0.49 | 6 | 0.07 | -0.08, 0.22 |
| | VAT | 128 | 0.24 | 0.2, 0.28 | 4 | 0.26 | 0.05, 0.46 |
| Moderate PA | LBM | 10 | 0.13 | -2.26, 2.52 | -- | -- | -- |
| | BMI | 21 | 0.11 | -0.11, 0.32 | -- | -- | -- |
| | WHR | 20 | 0.15 | -0.1, 0.4 | -- | -- | -- |
| | WC | 21 | 0.22 | -0.02, 0.46 | -- | -- | -- |
| | %TBF | 20 | -0.05 | -0.35, 0.25 | -- | -- | -- |
| | VAT | 32 | 0.03 | -0.05, 0.12 | -- | -- | -- |
| MVPA | LBM | 39 | 0.57 | -0.72, 1.86 | 14 | 0.25 | -0.9, 1.4 |
| | BMI | 72 | 0.06 | -0.06, 0.18 | 25 | -0.1 | -0.2, 0 |
| | WHR | 72 | 0.03 | -0.11, 0.16 | 26 | -0.03 | -0.15, 0.09 |
| | WC | 72 | -0.02 | -0.15, 0.1 | 26 | -0.06 | -0.18, 0.06 |
| | %TBF | 66 | -0.12 | -0.28, 0.05 | 24 | -0.15 | -0.3, -0.01 |
| | VAT | 93 | -0.07 | -0.12, -0.01 | 46 | -0.09 | -0.16, -0.03 |
| Vigorous PA | LBM | 36 | 0.22 | -2.43, 2.86 | 3 | -1.07 | -3.24, 1.09 |
| | BMI | 38 | -0.05 | -0.33, 0.23 | 15 | -0.12 | -0.25, 0.01 |
| | WHR | 38 | 0.09 | -0.22, 0.39 | 15 | -0.06 | -0.23, 0.11 |
| | WC | 40 | -0.2 | -0.49, 0.1 | 15 | -0.08 | -0.23, 0.06 |
| | %TBF | 33 | -0.07 | -0.44, 0.3 | 15 | -0.2 | -0.4, 0 |
| | VAT | 61 | -0.14 | -0.24, -0.04 | 26 | -0.19 | -0.28, -0.1 |
| SSOE | LBM | 19 | 0.64 | -2.57, 3.84 | -- | -- | -- |
| | BMI | 74 | -0.34 | -0.59, -0.1 | -- | -- | -- |
| | WHR | 74 | -0.12 | -0.38, 0.15 | -- | -- | -- |
| | WC | 77 | -0.06 | -0.31, 0.19 | -- | -- | -- |
| | %TBF | 72 | -0.26 | -0.59, 0.07 | -- | -- | -- |
| | VAT | 98 | -0.31 | -0.41, -0.21 | -- | -- | -- |
| Overall Activity | LBM | -- | -- | -- | 7 | 1.12 | -0.7, 2.93 |
| | BMI | -- | -- | -- | 4 | -0.18 | -0.43, 0.07 |
| | WHR | -- | -- | -- | 4 | -0.16 | -0.41, 0.1 |
| | WC | -- | -- | -- | 4 | -0.14 | -0.35, 0.07 |
| | %TBF | -- | -- | -- | 4 | -0.29 | -0.59, 0.02 |
| | VAT | -- | -- | -- | 6 | -0.21 | -0.38, -0.03 |

Abbreviations: MR, Mendelian randomization; SNPs, single nucleotide polymorphisms; PA, physical activity; MVPA, moderate-to-vigorous PA; SSOE, strenuous sports or other exercises; LBM, lean body mass; BMI, body mass index; WHR, waist-to-hip ratio; WC, waist circumference; %TBF, total body fat percent; VAT, visceral adipose tissue.

Note: The weighted median causal estimates are based on the relationship between the strength of association between the SNP and exposure, and the SNP and outcome associations.

**Table S7.** MR Mode-based Estimates for Physical Activity to Body Composition

| Phenotypes | | Self-Report MR | | | Accelerometer MR | | |
|---|---|---|---|---|---|---|---|
| *Activity* | *Body Comp.* | *SNPs (#)* | *beta* | *95% CI* | *SNPs (#)* | *beta* | *95% CI* |
| Sedentary Behavior | LBM | 55 | 0.38 | -1.53, 2.29 | 4 | 0.24 | 0.02, 0.46 |
| | BMI | 55 | 0.21 | -0.03, 0.44 | 4 | 0.37 | -0.08, 0.82 |
| | WHR | 56 | 0.23 | -0.01, 0.48 | 4 | 0.44 | 0.19, 0.69 |
| | WC | 59 | 0.16 | -0.08, 0.39 | 3 | -0.2 | -0.58, 0.18 |
| | %TBF | 53 | 0.46 | 0.17, 0.75 | 6 | 0.03 | -0.17, 0.24 |
| | VAT | 128 | 0.24 | 0.13, 0.36 | 4 | 0.24 | 0.02, 0.46 |
| Moderate PA | LBM | 10 | 0.4 | -3.55, 4.35 | -- | -- | -- |
| | BMI | 21 | 0.21 | -0.21, 0.64 | -- | -- | -- |
| | WHR | 20 | 0.14 | -0.36, 0.63 | -- | -- | -- |
| | WC | 21 | 0.2 | -0.29, 0.69 | -- | -- | -- |
| | %TBF | 20 | -0.19 | -0.75, 0.36 | -- | -- | -- |
| | VAT | 32 | 0.04 | -0.13, 0.21 | -- | -- | -- |
| MVPA | LBM | 39 | 0.62 | -2.2, 3.45 | 14 | 0.69 | -1.56, 2.93 |
| | BMI | 72 | -0.01 | -0.32, 0.29 | 25 | -0.1 | -0.27, 0.07 |
| | WHR | 72 | 0.1 | -0.24, 0.44 | 26 | -0.06 | -0.3, 0.18 |
| | WC | 72 | 0.05 | -0.23, 0.32 | 26 | -0.06 | -0.34, 0.23 |
| | %TBF | 66 | -0.2 | -0.57, 0.17 | 24 | -0.19 | -0.44, 0.06 |
| | VAT | 93 | -0.04 | -0.17, 0.09 | 46 | -0.03 | -0.18, 0.11 |
| Vigorous PA | LBM | 36 | 3.5 | -2.73, 9.74 | 3 | -1.73 | -4.91, 1.46 |
| | BMI | 38 | -0.15 | -0.86, 0.56 | 15 | -0.17 | -0.38, 0.03 |
| | WHR | 38 | 0.36 | -0.17, 0.88 | 15 | -0.05 | -0.32, 0.22 |
| | WC | 40 | 0.4 | -0.27, 1.07 | 15 | -0.09 | -0.35, 0.17 |
| | %TBF | 33 | 0.44 | -0.44, 1.32 | 15 | -0.27 | -0.57, 0.03 |
| | VAT | 61 | -0.09 | -0.34, 0.15 | 26 | -0.23 | -0.39, -0.06 |
| SSOE | LBM | 19 | 4.44 | -2.06, 10.93 | -- | -- | -- |
| | BMI | 74 | -0.51 | -1.18, 0.16 | -- | -- | -- |
| | WHR | 74 | 0.04 | -0.85, 0.93 | -- | -- | -- |
| | WC | 77 | 0.13 | -0.48, 0.74 | -- | -- | -- |
| | %TBF | 72 | 0.05 | -0.67, 0.78 | -- | -- | -- |
| | VAT | 98 | -0.37 | -0.73, 0 | -- | -- | -- |
| Overall Activity | LBM | -- | -- | -- | 7 | 2.56 | -0.86, 5.98 |
| | BMI | -- | -- | -- | 4 | -0.04 | -0.55, 0.46 |
| | WHR | -- | -- | -- | 4 | -0.04 | -0.34, 0.26 |
| | WC | -- | -- | -- | 4 | -0.28 | -0.61, 0.05 |
| | %TBF | -- | -- | -- | 4 | -0.26 | -0.67, 0.15 |
| | VAT | -- | -- | -- | 6 | -0.17 | -0.39, 0.06 |

Abbreviations: MR, Mendelian randomization; SNPs, single nucleotide polymorphisms; PA, physical activity; MVPA, moderate-to-vigorous PA; SSOE, strenuous sports or other exercises; LBM, lean body mass; BMI, body mass index; WHR, waist-to-hip ratio; WC, waist circumference; %TBF, total body fat percent; VAT, visceral adipose tissue.

Note: The weighted mode causal estimates are based on the relationship between the strength of association between the SNP and exposure, and the SNP and outcome associations. The weighted-mode estimate uses the property that valid instruments should provide the largest number of similar individual-instrument causal estimates even if the majority of instruments is invalid.

**Table S8.** MR-EGGER Estimates for Physical Activity to Body Composition

| **Phenotypes** | | **Self-Report MR** | | | **Accelerometer MR** | | |
|---|---|---|---|---|---|---|---|
| *Activity* | *Body Comp.* | *SNPs (#)* | *beta* | *95% CI* | *SNPs (#)* | *beta* | *95% CI* |
| Sedentary Behavior | LBM | 55 | 0 | -0.24, 0.24 | 4 | -0.25 | -1.24, 0.74 |
| | BMI | 55 | 0.53 | 0, 1.06 | 4 | -0.32 | -2.81, 2.17 |
| | WHR | 56 | 0.54 | 0.1, 0.99 | 4 | 0.33 | -3.35, 4.02 |
| | WC | 59 | 0.37 | -0.03, 0.77 | 3 | 0.14 | -3.02, 3.29 |
| | %TBF | 53 | 1.03 | 0.45, 1.62 | 6 | -1.87 | -4.7, 0.95 |
| | VAT | 128 | 0.34 | 0.13, 0.56 | 4 | 0 | -0.69, 0.69 |
| Moderate PA | LBM | 10 | -0.05 | -0.26, 0.15 | -- | -- | -- |
| | BMI | 21 | -0.36 | -1.23, 0.51 | -- | -- | -- |
| | WHR | 20 | -0.14 | -0.95, 0.67 | -- | -- | -- |
| | WC | 21 | 0.09 | -0.81, 0.99 | -- | -- | -- |
| | %TBF | 20 | 0.18 | -0.82, 1.18 | -- | -- | -- |
| | VAT | 32 | 0.08 | -0.21, 0.38 | -- | -- | -- |
| MVPA | LBM | 39 | -0.15 | -0.35, 0.05 | 14 | 0.03 | -0.08, 0.15 |
| | BMI | 72 | -0.08 | -0.57, 0.41 | 25 | 0.11 | -0.44, 0.66 |
| | WHR | 72 | -0.2 | -0.74, 0.35 | 26 | 0.1 | -0.44, 0.63 |
| | WC | 72 | 0.11 | -0.4, 0.61 | 26 | 0.1 | -0.33, 0.54 |
| | %TBF | 66 | -0.12 | -0.7, 0.46 | 24 | 0.18 | -0.32, 0.68 |
| | VAT | 93 | -0.18 | -0.4, 0.03 | 46 | -0.09 | -0.24, 0.06 |
| Vigorous PA | LBM | 36 | 0.35 | -0.05, 0.75 | 3 | 0.18 | -0.39, 0.75 |
| | BMI | 38 | 0.62 | -0.41, 1.65 | 15 | -0.26 | -1.31, 0.8 |
| | WHR | 38 | 0.55 | -0.44, 1.53 | 15 | -0.09 | -1.26, 1.09 |
| | WC | 40 | 0.81 | -0.08, 1.71 | 15 | -0.27 | -1.27, 0.74 |
| | %TBF | 33 | 0.05 | -1.07, 1.17 | 15 | -1.37 | -2.76, 0.03 |
| | VAT | 61 | 0.05 | -0.2, 0.3 | 26 | -0.24 | -0.51, 0.02 |
| SSOE | LBM | 19 | 0.01 | -0.3, 0.31 | -- | -- | -- |
| | BMI | 74 | 0.32 | -1.08, 1.72 | -- | -- | -- |
| | WHR | 74 | 0.2 | -1.13, 1.54 | -- | -- | -- |
| | WC | 77 | 0.1 | -0.98, 1.18 | -- | -- | -- |
| | %TBF | 72 | 0.01 | -1.41, 1.42 | -- | -- | -- |
| | VAT | 98 | -0.09 | -0.45, 0.26 | -- | -- | -- |
| Overall Activity | LBM | -- | -- | -- | 7 | 0.14 | -0.07, 0.35 |
| | BMI | -- | -- | -- | 4 | 4.15 | -2.54, 10.84 |
| | WHR | -- | -- | -- | 4 | 4.58 | -1.08, 10.24 |
| | WC | -- | -- | -- | 4 | 2.19 | -1.08, 5.46 |
| | %TBF | -- | -- | -- | 4 | 0.95 | -3.56, 5.47 |
| | VAT | -- | -- | -- | 6 | -0.25 | -1.08, 0.57 |

Abbreviations: MR, Mendelian randomization; SNPs, single nucleotide polymorphisms; PA, physical activity; MVPA, moderate-to-vigorous PA; SSOE, strenuous sports or other exercises; LBM, lean body mass; BMI, body mass index; WHR, waist-to-hip ratio; WC, waist circumference; %TBF, total body fat percent; VAT, visceral adipose tissue.

Note: MR-Egger regression assumes linearity and homogeneity in the associations between the genetic variants, risk factor, and outcome. Egger regression provides an estimate of the causal effect that is consistent asymptotically even if all the genetic variants have pleiotropic effects on the outcome.

**Table S9.** CAUSE estimates for Physical Activity to Body Composition

| **Phenotypes** | | **Self-Report MR** | | **Accelerometer MR** | |
|---|---|---|---|---|---|
| *Activity* | *Body Comp.* | *Effect* | *95% CI* | *Effect* | *95% CI* |
| Sedentary Behavior | LBM | 0.00 | -0.36, 0.40 | 0.42 | -1.04, 1.85 |
| | BMI | 0.14 | 0.10, 0.18** | 0.05 | -0.06, 0.17 |
| | WHR | 0.13 | 0.08, 0.17** | 0.02 | -0.12, 0.17 |
| | WC | 0.16 | 0.11, 0.20** | 0.03 | -0.15, 0.20 |
| | %TBF | 0.17 | 0.11, 0.24 | 0.12 | -0.08, 0.33 |
| | VAT | 0.23 | 0.19, 0.28*** | 0.10 | -0.01, 0.21 |
| Moderate PA | LBM | -0.69 | -2.18, 0.81 | -- | -- |
| | BMI | 0.07 | -0.11, 0.26 | -- | -- |
| | WHR | 0.20 | 0.02, 0.39 | -- | -- |
| | WC | 0.13 | -0.10, 0.36 | -- | -- |
| | %TBF | 0.04 | -0.24, 0.32 | -- | -- |
| | VAT | 0.25 | 0.02, 0.47 | -- | -- |
| MVPA | LBM | 0.51 | -0.15, 1.24 | 0.00 | -0.47, 0.47 |
| | BMI | -0.02 | -0.11, 0.07 | -0.13 | -0.19, -0.07 |
| | WHR | -0.03 | -0.11, 0.06 | -0.08 | -0.15, -0.02 |
| | WC | -0.03 | -0.12, 0.07 | -0.12 | -0.19, -0.05 |
| | %TBF | -0.08 | -0.20, 0.03 | -0.11 | -0.19, -0.03 |
| | VAT | -0.06 | -0.16, 0.04 | -0.11 | -0.15, -0.07*** |
| Vigorous PA | LBM | 0.59 | -0.84, 2.05 | 0.15 | -0.42, 0.75 |
| | BMI | 0.00 | -0.18, 0.19 | -0.08 | -0.16, 0.01 |
| | WHR | -0.03 | -0.20, 0.14 | -0.02 | -0.10, 0.06 |
| | WC | 0.02 | -0.16, 0.21 | -0.07 | -0.16, 0.02 |
| | %TBF | -0.11 | -0.34, 0.12 | -0.07 | -0.19, 0.05 |
| | VAT | -0.13 | -0.33, 0.06 | -0.14 | -0.19, -0.08*** |
| SSOE | LBM | 1.36 | -0.19, 2.82 | -- | -- |
| | BMI | -0.19 | -0.36, -0.02 | -- | -- |
| | WHR | -0.22 | -0.37, -0.08 | -- | -- |
| | WC | -0.22 | -0.39, -0.05 | -- | -- |
| | %TBF | -0.34 | -0.56, -0.11 | -- | -- |
| | VAT | -0.28 | -0.46, -0.11 | -- | -- |
| Overall Activity | LBM | -- | -- | 0.18 | -0.98, 1.31 |
| | BMI | -- | -- | -0.12 | -0.28, 0.04 |
| | WHR | -- | -- | -0.09 | -0.25, 0.08 |
| | WC | -- | -- | -0.11 | -0.30, 0.07 |
| | %TBF | -- | -- | -0.17 | -0.35, 0.02 |
| | VAT | -- | -- | -0.08 | -0.19, 0.03 |

Abbreviations: MR, Mendelian randomization; SNPs, single nucleotide polymorphisms; PA, physical activity; MVPA, moderate-to-vigorous PA; SSOE, strenuous sports or other exercises; LBM, lean body mass; BMI, body mass index; WHR, waist-to-hip ratio; WC, waist circumference; %TBF, total body fat percent; VAT, visceral adipose tissue. Note: ***p<.001, **p<.01

**Table S10.** Bidirectional MR IVW Estimates for Body Composition to Physical Activity

| **Phenotypes** | | **Self-Report MR** | | | **Accelerometer MR** | | |
|---|---|---|---|---|---|---|---|
| *Body Comp.* | *Activity* | *SNPs (#)* | *beta* | *95% CI* | *SNPs (#)* | *beta* | *95% CI* |
| LBM† | Sedentary Behavior | 15 | 0.00 | -0.00, 0.00 | -- | -- | -- |
| | Moderate PA | 16 | 0.00 | -0.01, 0.00 | -- | -- | -- |
| | MVPA | 16 | 0.00 | -0.00, 0.01 | 16 | 0.00 | 0.00, 0.01 |
| | Vigorous PA | 16 | 0.00 | -0.00, 0.00 | 16 | 0.01 | 0.00, 0.02 |
| | SSOE | 16 | 0.00 | -0.00, 0.00 | -- | -- | -- |
| | Overall Activity | -- | -- | -- | -- | -- | -- |
| BMI | Sedentary Behavior | 67 | 0.09 | 0.08, 0.10 | 11 | 0.12 | -0.03, 0.26 |
| | Moderate PA | 78 | 0.01 | -0.02, 0.03 | 12 | -0.04 | -0.16, 0.07 |
| | MVPA | 78 | -0.01 | -0.04, 0.03 | 71 | -0.03 | -0.08, 0.02 |
| | Vigorous PA | 78 | 0.01 | -0.01, 0.03 | 71 | -0.08 | -0.13, -0.04 |
| | SSOE | 78 | 0.00 | -0.02, 0.02 | -- | -- | -- |
| | Overall Activity | -- | -- | -- | 11 | -0.03 | -0.17, 0.11 |
| %TBF | Sedentary Behavior | 6 | 0.12 | -0.04, 0.29 | 2 | 0.11 | -0.18, 0.40 |
| | Moderate PA | 9 | -0.05 | -0.10, 0.00 | 2 | 0.07 | -0.16, 0.31 |
| | MVPA | 9 | -0.05 | -0.17, 0.05 | 9 | -0.02 | -0.14, 0.10 |
| | Vigorous PA | 9 | -0.00 | -0.06, 0.05 | 9 | -0.00 | -0.11, 0.10 |
| | SSOE | 9 | 0.01 | -0.03, 0.04 | -- | -- | -- |
| | Overall Activity | -- | -- | -- | 2 | 0.07 | -0.3, 0.45 |
| VAT | Sedentary Behavior | 18 | 0.05 | -0.01, 0.11 | 28 | 0.23 | 0.10, 0.36 |
| | Moderate PA | 19 | -0.01 | -0.05, 0.03 | 29 | -0.12 | -0.25, 0.01 |
| | MVPA | 17 | 0.02 | -0.03, 0.07 | 236 | -0.17 | -0.22, -0.12 |
| | Vigorous PA | 17 | 0.03 | 0.00, 0.06 | 211 | -0.19 | -0.23, -0.14 |
| | SSOE | 17 | 0.03 | 0.02, 0.05 | -- | -- | -- |
| | Overall Activity | -- | -- | -- | 27 | -0.10 | -0.25, 0.05 |

Abbreviations: MR, Mendelian randomization; SNPs, single nucleotide polymorphisms; LBM, lean body mass; BMI, body mass index; %TBF, total body fat percent; VAT, visceral adipose tissue; PA, physical activity; MVPA, moderate-to-vigorous PA; SSOE, strenuous sports or other exercises.

†LBM contained one SNPs at the genome-wide significance level; a reduced IV threshold ($p < 5x10^{-6}$) was used for analyses. Outcome SNPs could not be identified in GWAS of sedentary behavior, moderate PA, or overall activity.

| SNP ID | Chromosome | Position | Gene | Allele | EAF | B | SE | *p* |
|---|---|---|---|---|---|---|---|---|
| rs6545389 | 2 | 54386901 | ACYP2 | A / G | 0.4997 | -0.0127 | 0.0023 | 3.5e-08 |

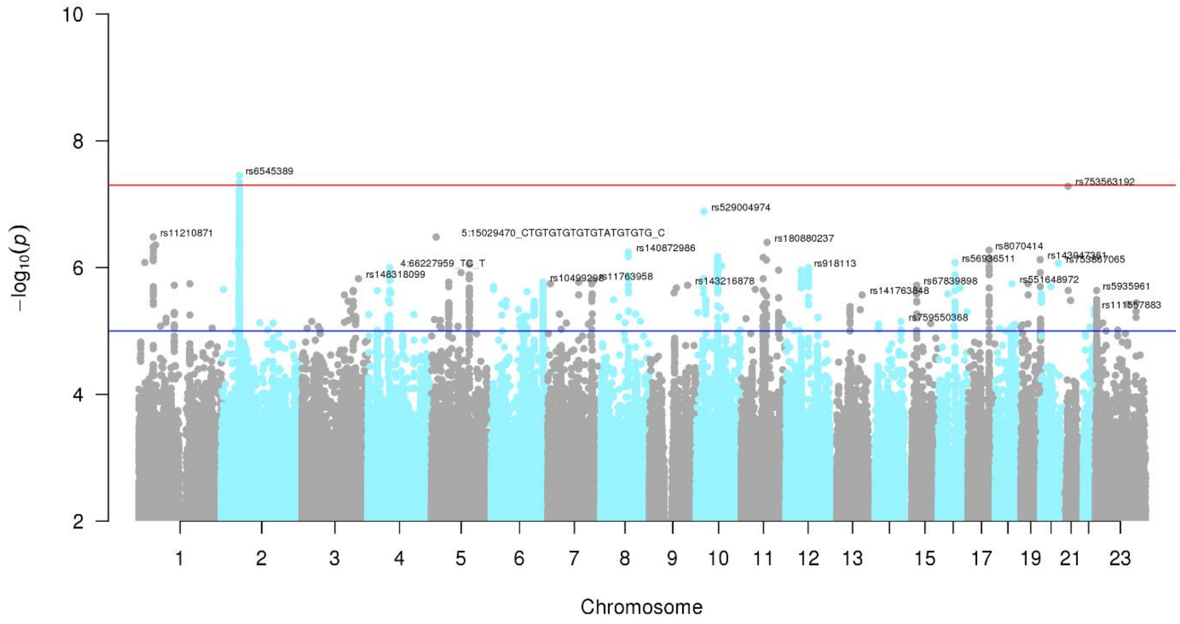

**Figure S1.** Manhattan Plot and genome-wide significant SNPs for self-reported moderate PA

| SNP ID | Chromosome | Position | Gene | Allele | EAF | B | SE | *p* |
|---|---|---|---|---|---|---|---|---|
| rs4480415 | 1 | 174800121 | RABGAP1L | G / A | 0.7248 | -0.012 | 0.0023 | 4.6e-08 |
| rs17443704 | 5 | 151668231 | -- | G / A | 0.8231 | -0.013 | 0.0023 | 3.5e-08 |

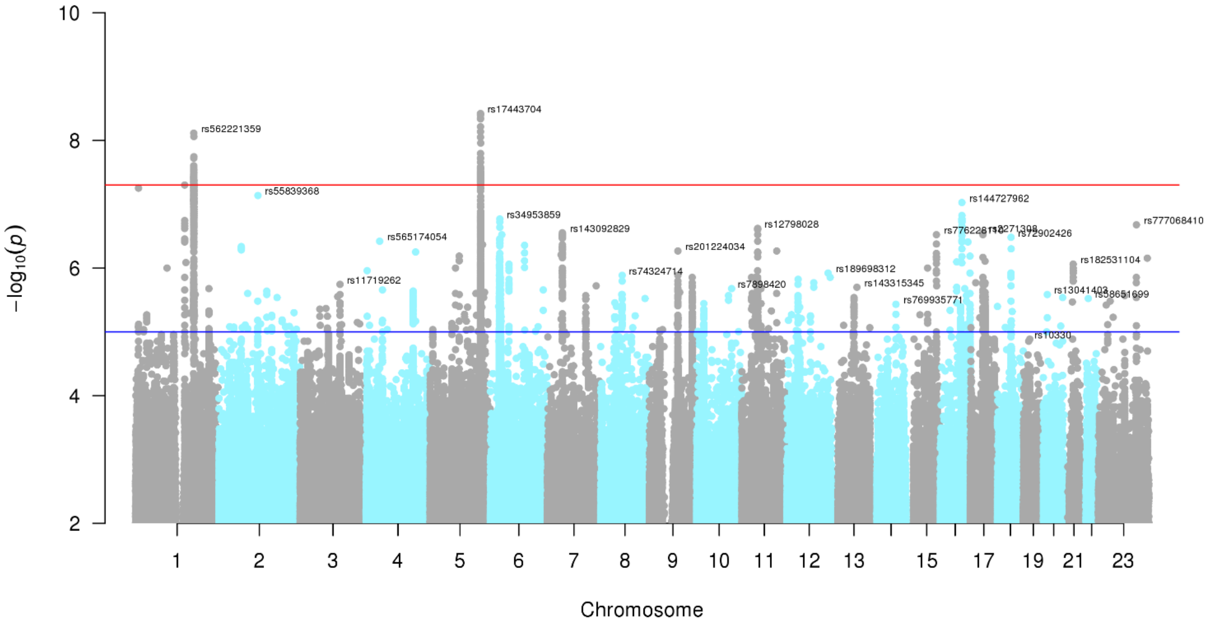

**Figure S2.** Manhattan Plot and genome-wide significant SNPs for accelerometer-measured MVPA

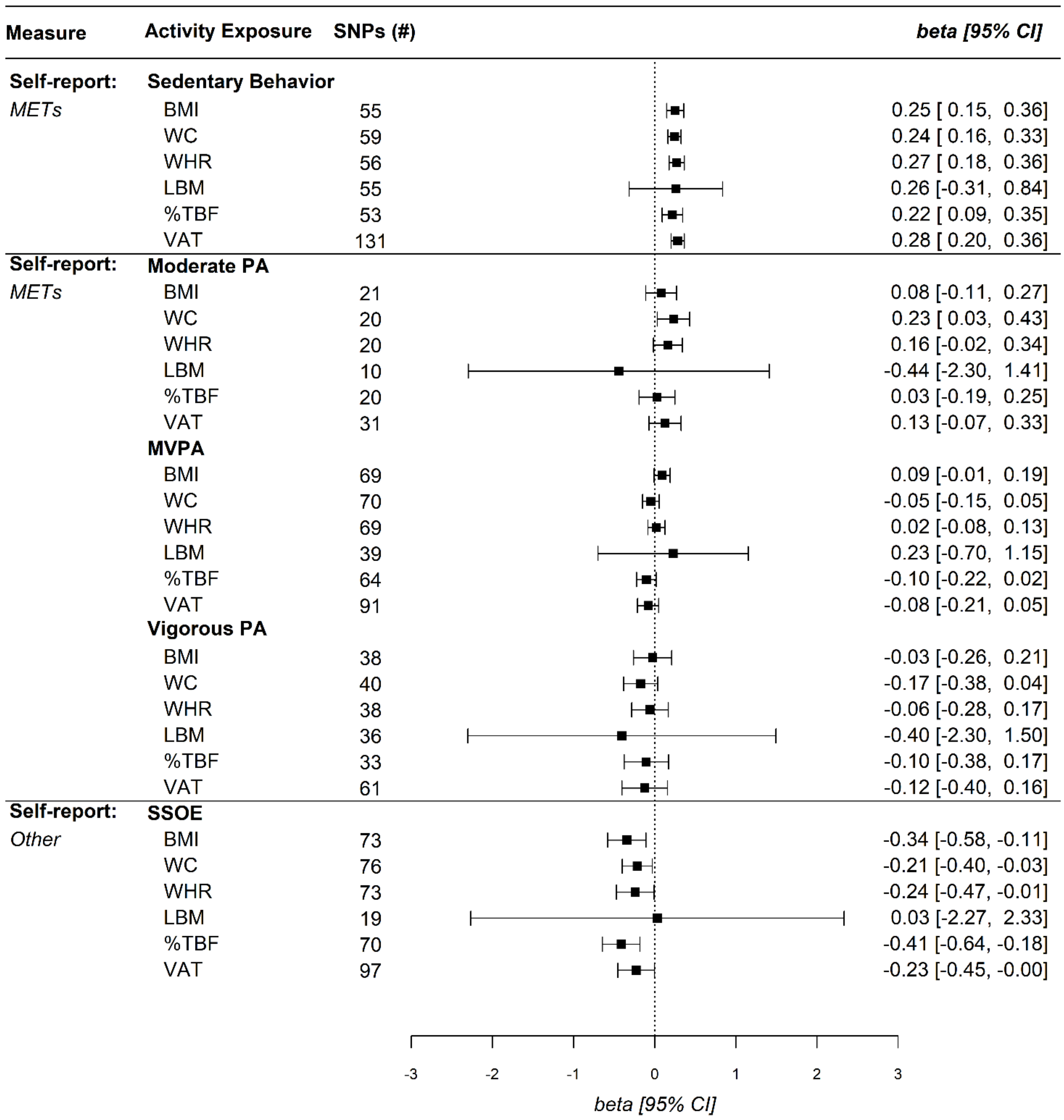

**Figure S3.** Univariable Mendelian Randomization IVW Sensitivity Analysis for Self-Reported Physical Activity Levels and Body Composition removing the *APOE* rs429358 variant. Abbreviations: SNPs, single nucleotide polymorphisms; CI, confidence interval; METs, metabolic equivalent of tasks; BMI, body mass index; WC, waist circumference; WHR, waist-to-hip ratio; LBM, lean body mass; BF %, total body fat percent; VAT, visceral adipose tissue; MVPA, moderate-to-vigorous PA; SSOE, strenuous sports or other exercises.

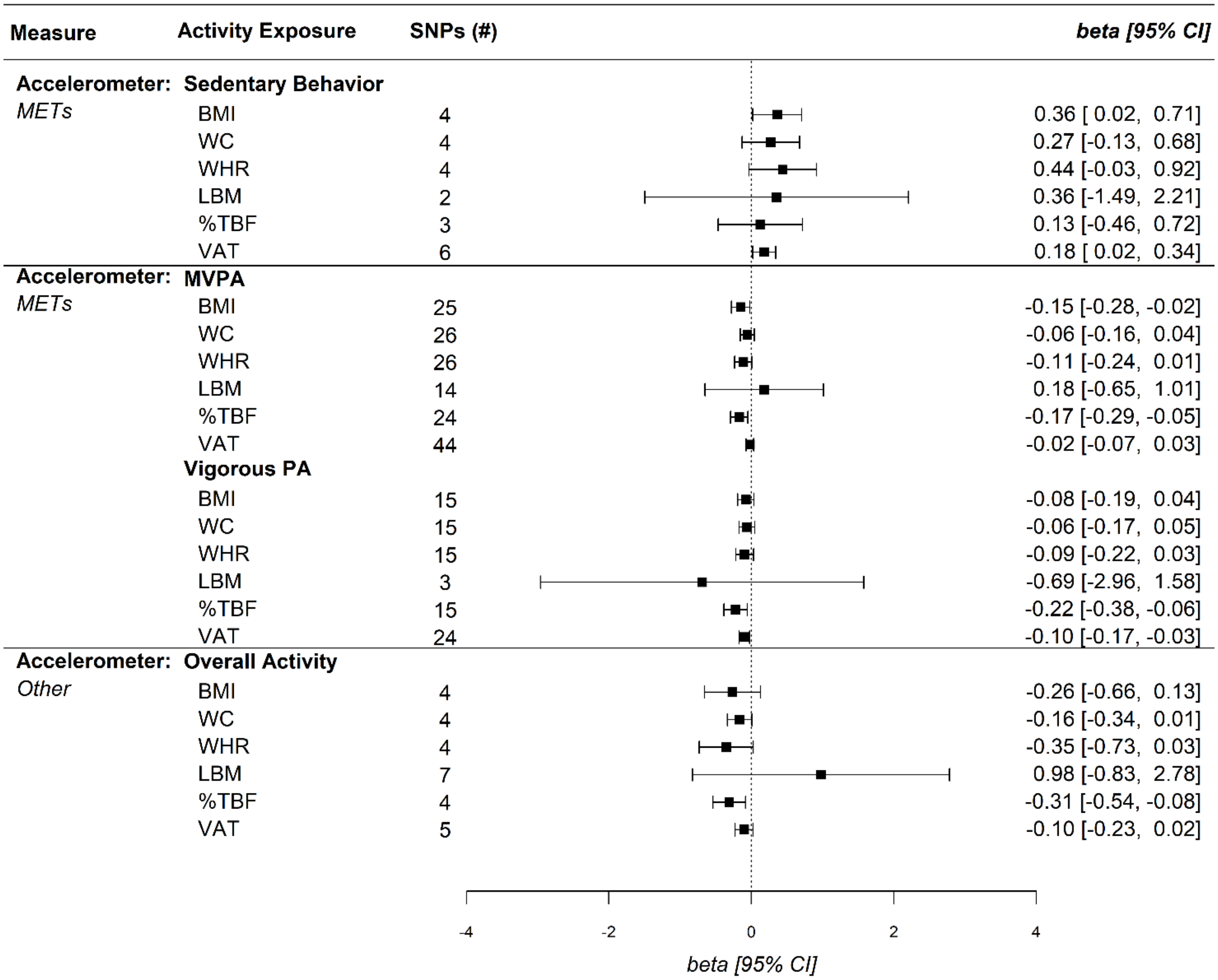

**Figure S4.** Univariable Mendelian Randomization IVW Sensitivity Analysis for Self-Reported Physical Activity Levels and Body Composition removing the *APOE* rs429358 variant. Abbreviations: SNPs, single nucleotide polymorphisms; CI, confidence interval; METs, metabolic equivalent of tasks; BMI, body mass index; WC, waist circumference; WHR, waist-to-hip ratio; LBM, lean body mass; BF %, total body fat percent; VAT, visceral adipose tissue; MVPA, moderate-to-vigorous PA.
